# Within-mouth spillover of dental restorations on periodontal status

**DOI:** 10.64898/2026.08.13.26360345

**Authors:** Sebastian-Edgar Baumeister, Daniel Hagenfeld, Michael Nolde, Stefanie Samietz, Philipp Kanzow, Henry Völzke, Thomas Kocher, Birte Holtfreter

**Author notes:** Corresponding author: Sebastian-Edgar Baumeister, Institute of Health Services Research in Dentistry, University of Münster, Albert-Schweitzer-Campus 1, 48149 Münster, Germany.

## Abstract

**Aim:** To test whether neighbouring-tooth restorations affect a focal tooth’s periodontal status (within-mouth spillover), and whether accounting for them alters the established association at the restored surface.

**Materials and methods:** Prospective tooth-surface data from the Study of Health in Pomerania (SHIP): SHIP-START (N=2,715/1,982/1,397, follow-ups 1–3) and SHIP-TREND (N=2,123). Own and neighbouring-tooth restoration were defined at baseline; probing depth (PD), clinical attachment level (CAL), bleeding on probing (BOP) and deep pockets (PD ≥4 mm) at follow-up. A neighbourhood-exposure mixed model with generalized propensity-score adjustment estimated both effects, with two corroborating estimators.

**Results:** Own restoration was associated with worse periodontal status at that surface (crown PD exp(β) up to 1.10). Neighbouring-tooth restorations raised focal-tooth PD (spillover exp(β) 1.03–1.05) and deep-pocket risk (risk ratio up to 1.25); CAL and BOP showed none consistently. It attenuated with tooth-lag, nearing the null by two to three teeth in most estimators. Ignoring interference modestly inflated the direct effect; an opposing-tooth negative control showed none; untreated caries reproduced it.

**Conclusions:** Dental restorations exert a within-mouth spillover on neighbouring-tooth PD: their periodontal footprint extends beyond the restored tooth. The direct association persists, modestly attenuated, after accounting for the neighbourhood.

**Clinical Relevance:** *Scientific rationale for the study:* Conventional surface-level analyses treat teeth as independent (the no-interference assumption) and cannot detect whether a tooth’s restoration affects the periodontal status of neighbouring teeth.

*Principal findings:* Restorations on neighbouring teeth independently raised focal-tooth PD and deep-pocket risk; the spillover was local, specific to PD, and concordant for untreated caries. Ignoring interference inflated the direct own-restoration effect only modestly.

*Practical implications:* A restoration’s periodontal footprint extends modestly beyond the restored surface to the neighbouring tooth; the dominant effect nonetheless remains local, and own-surface estimates overstate the direct effect only modestly.

## Introduction

Severe periodontitis affected more than a billion people in 2021 (Nascimento et al. 2024) and co-leads the oral-disease burden with dental caries, with which it shares determinants (Baima et al. 2023; Romandini et al. 2024; Trindade et al. 2023). Rising tooth retention brings more restored teeth into the ages at which periodontitis concentrates (Kocher et al. 2025).

Restorations have long been associated with worse periodontal status at the treated site: overhanging and subgingival margins promote biofilm retention and gingival inflammation and can impinge on the supracrestal tissue attachment (Gargiulo et al. 1961; Gracis et al. 2023). Subgingival margins caused attachment loss within one to three years (Schätzle et al. 2001) and overhangs deeper pockets and interproximal bone loss (Jansson et al. 1994; Millar and Blake 2019); in the Dunedin cohort an interproximal caries or restorative event roughly doubled the odds of ≥3 mm attachment loss at the neighbouring site (Broadbent et al. 2006), and an adolescent study found breakdown at the neighbouring tooth’s approximal surface (Albandar et al. 1995). In SHIP-TREND, a cohort of the Study of Health in Pomerania (SHIP), crowned surfaces carried a 41% higher risk of bleeding on probing (BOP; risk ratio, RR, 1.41 [1.33, 1.50]; Nafz et al. 2026), and a German survey reported probing depth (PD) 0.16–0.37 mm higher at restored sites (Pitchika et al. 2026).

All of this evidence estimates the effect of a focal surface’s own restoration on the periodontal site at that surface, treating teeth as independent units and invoking the stable unit treatment value assumption (SUTVA; Rubin 1980): that one tooth’s restoration does not affect another tooth’s periodontium. Models conditioning only on a surface’s own restoration thus ignore restorations elsewhere; the mouth is not a collection of independent units.

It is a tightly packed spatial system: neighbouring teeth share interproximal contacts, the papilla and a continuous supragingival biofilm, forming a common ecological niche, so a subgingival or overhanging margin may alter the local ecology of the neighbouring tooth. In causal-inference terms this is interference, or spillover: a focal tooth’s periodontal status may depend not only on its own restoration (the direct effect) but also on neighbouring-tooth restorations (the indirect, or spillover, effect; Figure 1a). Because interference operates within but not between mouths, the mouth is the natural cluster for a partial-interference analysis (Hudgens and Halloran 2008; Tchetgen Tchetgen and VanderWeele 2012; Figure 1b). If spillover exists, the periodontal burden of restorations is a whole-mouth quantity that own-surface estimates understate.

**Figure 1.**
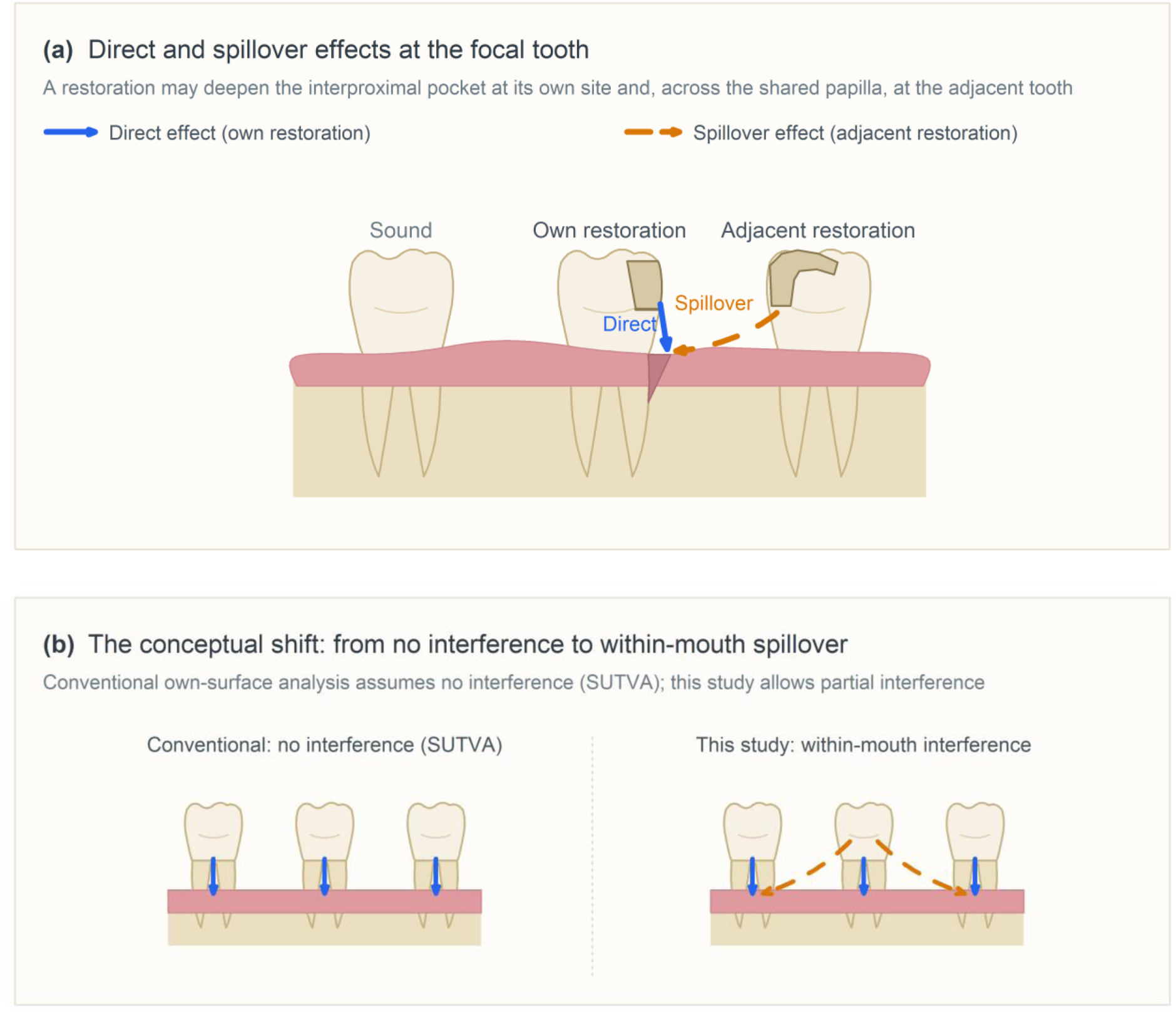
Conceptual illustration of within-mouth interference of dental restorations on periodontal status. (a) Direct and spillover effects at a focal tooth. The focal tooth and the neighbouring tooth each carry a proximal restoration on the coronal half of the interproximal surface, and a deep periodontal pocket has formed in the shared interdental space. The focal tooth’s own restoration may deepen the pocket at its own site (direct effect; solid arrow), while the restoration on the neighbouring tooth may act across the shared interdental papilla to deepen the same pocket (spillover, or indirect, effect; dashed arrow); both effects act on the soft-tissue attachment, not directly on bone. (b) The conceptual shift underlying this study: conventional analyses assume each restoration affects only its own site (no interference; SUTVA), whereas the present study allows restorations to affect neighbouring teeth within the mouth (partial interference).

We therefore quantified within-mouth interference of dental restorations on periodontal status. With surface-level data from two population-based cohorts (SHIP-START, primary; SHIP-TREND, replication), we estimated the direct effect of a focal surface’s own restoration and the spillover effect of neighbouring-tooth restorations on PD, clinical attachment level (CAL), BOP and deep pockets, combining a neighbourhood-exposure mixed model with generalized propensity-score adjustment (Forastiere et al. 2022), a joint correlated-random-effects model (McNealis et al. 2026) and a doubly-robust estimator for partial interference (Qu et al. 2026), and repeating the analysis for untreated caries.

## Material and methods

### Study design and data

We used data from SHIP, a population-based research project in West Pomerania, north-east Germany (Völzke et al. 2022). For each cohort a stratified random sample of adults aged 20–79 years was drawn from local population registries. SHIP-START examined 4,307 participants at baseline (SHIP-START-0, 1997–2001; response 68.8%) and re-examined them at SHIP-START-1 (2002–2006), SHIP-START-2 (2008–2012) and SHIP-START-3 (2014–2016). SHIP-TREND examined 4,420 adults at baseline (SHIP-TREND-0, 2008– 2012; response 50.1%) and re-examined them at SHIP-TREND-1 (2014–2018). Outcomes were assessed a mean of 4.7 (SD 0.7), 10.6 (0.8) and 15.3 (0.7) years after baseline at SHIP-START-1, -2 and -3, and 7.4 (0.8) years at SHIP-TREND-1.

The design is prospective: restoration status and covariates were measured at baseline and periodontal outcomes at follow-up. After exclusion of edentulous participants, 2,715 persons at SHIP-START-1, 1,982 at SHIP-START-2, 1,397 at SHIP-START-3 and 2,123 at SHIP-TREND-1 had dental-examination data entering the prepared datasets; excluding records with missing exposure or covariate data, the complete-case estimation samples comprised 2,584, 1,903, 1,353 and 2,066 persons (Supplementary Table S1; participants retained versus lost to each follow-up are compared in Supplementary Table S2), each contributing up to four surfaces per tooth (the distal, buccal, mesial and palatal/lingual probing sites); occlusal surfaces, which do not border the gingival margin, and third molars were excluded.

### Unit of analysis, exposures and outcomes

The unit of analysis was the person × tooth × surface. At baseline each focal surface was assigned a binary own exposure — restoration (filled or crowned) versus sound — following Nafz et al. (2026); untreated caries was analysed analogously as a disjoint confirmatory exposure. Probing sites were mapped to caries/restoration surfaces as in Nafz et al. (2026): distobuccal, midbuccal, mesiobuccal and midlingual/midpalatinal sites to the distal, buccal, mesial and palatal/lingual surfaces of the same tooth. The neighbourhood of a focal tooth comprised the teeth one position mesial and one distal within the quadrant (positions ±1); the neighbourhood exposure AN(i) is their mean exposure, and spillover coefficients contrast full neighbourhood exposure (AN(i)=1) with none (AN(i)=0). Four follow-up outcomes were analysed: PD and CAL in millimetres, BOP, and a deep pocket (PD ≥4 mm); BOP was recorded at index teeth only, so those analyses rest on a smaller, non-random subset of teeth. Examination procedures and full definitions are in the Supplement (Section A).

### Covariates

Confounders were selected by the modified disjunctive cause criterion (VanderWeele 2019), measured at baseline and harmonised across cohorts — age, sex, school education, smoking status, pack-years, body mass index, diabetes, toothbrushing frequency, interdental care, dental visit within 12 months, and probing site (Supplement, Section A). Restored surfaces were more prevalent among older participants and more frequent dental attenders; inverse-probability-of-treatment weighting reduced the maximum standardised mean difference from 0.27 to 0.03 in SHIP-START and from 0.48 (age) to 0.08 in SHIP-TREND (Supplementary Tables S3–S4).

### Statistical analysis

We distinguish the direct effect — of the focal surface’s restoration on the periodontal site at that surface — from the spillover effect of neighbouring-tooth restorations on the focal site; a non-zero spillover is direct evidence against the no-interference assumption. All models adjusted for the same baseline confounder set and the baseline outcome, and each SHIP-START follow-up was analysed in a separate model against the common baseline. We designate the PD spillover in SHIP-START under the generalized-propensity-score model as the primary analysis; other outcomes, waves, estimators and the untreated-caries exposure are secondary or confirmatory.

We estimated the direct effect with a generalized linear mixed model (GLMM): Gamma log-link for PD and CAL and Poisson log-link for the binary outcomes, with random intercepts for person, jaw and tooth, fitted by integrated nested Laplace approximation (R-INLA). Model A contrasted baseline restoration category (filling; full or partial crown) with sound; Model B, restricted to baseline-sound surfaces, estimated incident restoration.

The primary spillover analysis followed the neighbourhood-interference framework of Forastiere et al. (2022). A three-level model with the same random intercepts and links, including own and neighbourhood exposure, estimated the direct and spillover effects simultaneously; under no interference the neighbourhood coefficient is zero. Against confounding of the two exposures we adjusted for the individual propensity φ(X) = P(own restored | X) and the neighbourhood generalized propensity score λ(g; z, X), both entered through natural splines. The PD spillover agreed with exact Markov-chain Monte Carlo (nutpie) to three decimal places. Model and propensity scores are specified in the Supplement (Section B, equations S1–S3).

Two further estimators corroborated the spillover. The joint correlated-random-effects model of McNealis et al. (2026) models outcome and own exposure jointly with correlated mouth-level random effects, separating local spillover from shared mouth-level confounding; the indirect effect came from a Bernoulli allocation contrast (neighbourhood prevalence 0.7 vs. 0.3; equations S4–S6). The augmented inverse-probability-weighted (AIPW) estimator of Qu et al. (2026), defined over the joint exposure (own restoration, any restored neighbouring tooth), returns the average spillover effect in millimetres (equations S7–S8). These models were fitted by R-INLA; the joint model’s binary outcomes did not converge under INLA and were checked by exact Markov-chain Monte Carlo (nutpie), not tabulated. Bayesian estimates are posterior medians with 95% credible intervals (CrI); the doubly-robust AIPW estimator is frequentist and reported with 95% confidence intervals (CI).

We re-estimated the neighbourhood effect by topological tooth-lag d — d = 1 for the neighbouring tooth and d = 2, 3 for teeth two and three positions away within the quadrant — motivated by the spatial-treatment distance-decay framework of Pollmann (2026). Each estimator was refitted with the lag-d exposure, the mixed and propensity-score models adjusting for lags 1–3 simultaneously; the covariate-adjusted mixed model including the lag- 1–3 exposures jointly without propensity-score adjustment is denoted the mutual GLMM.

For binary outcomes the spillover was checked across logit, linear-probability and probit links (Supplementary Table S5). Positivity, within-person spatial autocorrelation (Moran’s I before and after neighbourhood adjustment) and identification (the intraclass correlation of the neighbourhood exposure) were examined (Debarsy and Le Gallo 2025; Reich et al. 2021; Supplementary Table S6). In sensitivity analyses we additionally adjusted for tooth type; stratified by approximal and smooth focal surfaces; redefined the neighbourhood exposure as the facing surface of the neighbouring tooth; repeated the analysis with incident restorations; and estimated a negative-control ‘spillover’ from the opposing (antagonist) tooth, which shares person-, side- and examiner-level confounding but no interproximal contact (Supplementary Section I).

Analyses used R 4.6.0 (R-INLA, data.table) and Stan 2.39.0 (nutpie). Reporting followed STROBE.

## Results

### Descriptive characteristics

Subject-level characteristics are given in Supplementary Table S1; weighting achieved good covariate balance (Supplementary Tables S3–S4).

### Direct effect of own restoration

A focal surface’s own restoration was associated with worse periodontal status at that surface (Table 1). Crowns showed the largest direct effects on PD (exp(β) 1.069 [1.061, 1.078] at SHIP-START-1, 1.096 [1.083, 1.109] at SHIP-START-3 and 1.091 [1.083, 1.100] in SHIP-TREND), with concordant increases in CAL (crown exp(β) up to 1.293 [1.163, 1.438]), BOP (RR up to 1.41 [1.32, 1.50]) and deep pockets (PD ≥4 mm; RR up to 1.59 [1.44, 1.75]); fillings showed smaller but consistent increases. Among baseline-sound surfaces, incident restorations reproduced the pattern (Model B, resting on 5,570/7,076/6,276/5,024 incident restored surfaces; incident crown PD exp(β) up to 1.115 [1.101, 1.129]).

**Table 1.** Direct effect of restoration status on periodontal status at the restored surface (Model A: baseline status; Model B: incident restoration). GLMM (Gamma-log for PD and CAL; Poisson-log for BOP and PD ≥4 mm; person/jaw/tooth random intercepts); exp(β) for PD and CAL, RRs for BOP and PD ≥4 mm, with 95% CrI.

| Model | Outcome | Category | START-1 | START-2 | START-3 | TREND-1 |
| --- | --- | --- | --- | --- | --- | --- |
| A: baseline status | BOP | Crown | 1.185 [1.112, 1.263] | 1.150 [1.071, 1.235] | 1.300 [1.194, 1.416] | 1.405 [1.318, 1.497] |
| A: baseline status | BOP | Filling | 1.079 [1.019, 1.143] | 1.123 [1.056, 1.194] | 1.187 [1.103, 1.277] | 1.162 [1.087, 1.242] |
| A: baseline status | CAL | Crown | 1.196 [1.045, 1.370] | 1.293 [1.163, 1.438] | 1.224 [1.090, 1.375] | 1.182 [1.106, 1.263] |
| A: baseline status | CAL | Filling | 1.078 [1.059, 1.098] | 1.094 [1.076, 1.112] | 1.068 [1.047, 1.090] | 1.030 [1.017, 1.043] |
| A: baseline status | PD | Crown | 1.069 [1.061, 1.078] | 1.094 [1.084, 1.104] | 1.096 [1.083, 1.109] | 1.091 [1.083, 1.100] |
| A: baseline status | PD | Filling | 1.022 [1.016, 1.028] | 1.030 [1.024, 1.037] | 1.034 [1.025, 1.042] | 1.015 [1.009, 1.021] |
| A: baseline status | PD $\geq 4$ mm | Crown | 1.223 [1.153, 1.298] | 1.453 [1.355, 1.558] | 1.585 [1.438, 1.747] | 1.509 [1.410, 1.614] |
| A: baseline status | PD $\geq 4$ mm | Filling | 1.142 [1.087, 1.200] | 1.244 [1.174, 1.317] | 1.366 [1.261, 1.480] | 1.185 [1.111, 1.263] |
| B: incident | BOP | Crown (incident) | 1.363 [1.192, 1.560] | 1.368 [1.225, 1.529] | 1.554 [1.386, 1.743] | 1.486 [1.267, 1.742] |
| B: incident | BOP | Filling (incident) | 1.077 [0.955, 1.214] | 1.060 [0.953, 1.180] | 1.217 [1.083, 1.368] | 1.240 [1.097, 1.403] |
| B: incident | CAL | Crown (incident) | 1.111 [0.881, 1.402] | 1.388 [1.222, 1.578] | 1.194 [1.038, 1.373] | 1.172 [1.040, 1.320] |
| B: incident | CAL | Filling (incident) | 1.078 [1.041, 1.115] | 1.071 [1.045, 1.097] | 1.057 [1.029, 1.086] | 1.038 [1.015, 1.061] |
| B: incident | PD | Crown (incident) | 1.054 [1.038, 1.071] | 1.115 [1.101, 1.129] | 1.112 [1.096, 1.128] | 1.095 [1.078, 1.112] |
| B: incident | PD | Filling (incident) | 1.027 [1.015, 1.038] | 1.015 [1.005, 1.025] | 1.033 [1.020, 1.046] | 1.026 [1.015, 1.038] |
| B: incident | PD $\geq 4$ mm | Crown (incident) | 1.068 [0.944, 1.208] | 1.351 [1.211, 1.508] | 1.489 [1.304, 1.700] | 1.315 [1.117, 1.548] |
| B: incident | PD $\geq 4$ mm | Filling (incident) | 1.169 [1.055, 1.295] | 1.294 [1.176, 1.425] | 1.441 [1.270, 1.634] | 1.179 [1.041, 1.336] |

### Spillover from neighbouring restorations

Under the generalized propensity score, restoration of neighbouring teeth independently raised focal-tooth PD beyond the tooth’s own restoration (spillover exp(β) 1.026 [1.019, 1.033] at SHIP-START-1, rising to 1.044 [1.034, 1.054] at SHIP-START-3 and 1.053 [1.046, 1.060] in SHIP-TREND) and raised deep-pocket risk (PD ≥4 mm RR 1.107 [1.045, 1.172] to 1.245 [1.133, 1.368]). CAL and BOP showed no consistent spillover; two intervals excluded 1 in the protective direction (CAL exp(β) 0.984 [0.970, 0.999] in SHIP-TREND; BOP RR 0.912 [0.847, 0.983] at SHIP-START-2) (Figure 2; Supplementary Table S7). The own-restoration (direct) effect remained positive in the same model but was attenuated (PD 1.017–1.027) relative to the otherwise identical model without the neighbourhood term (1.026–1.035).

**Figure 2.**
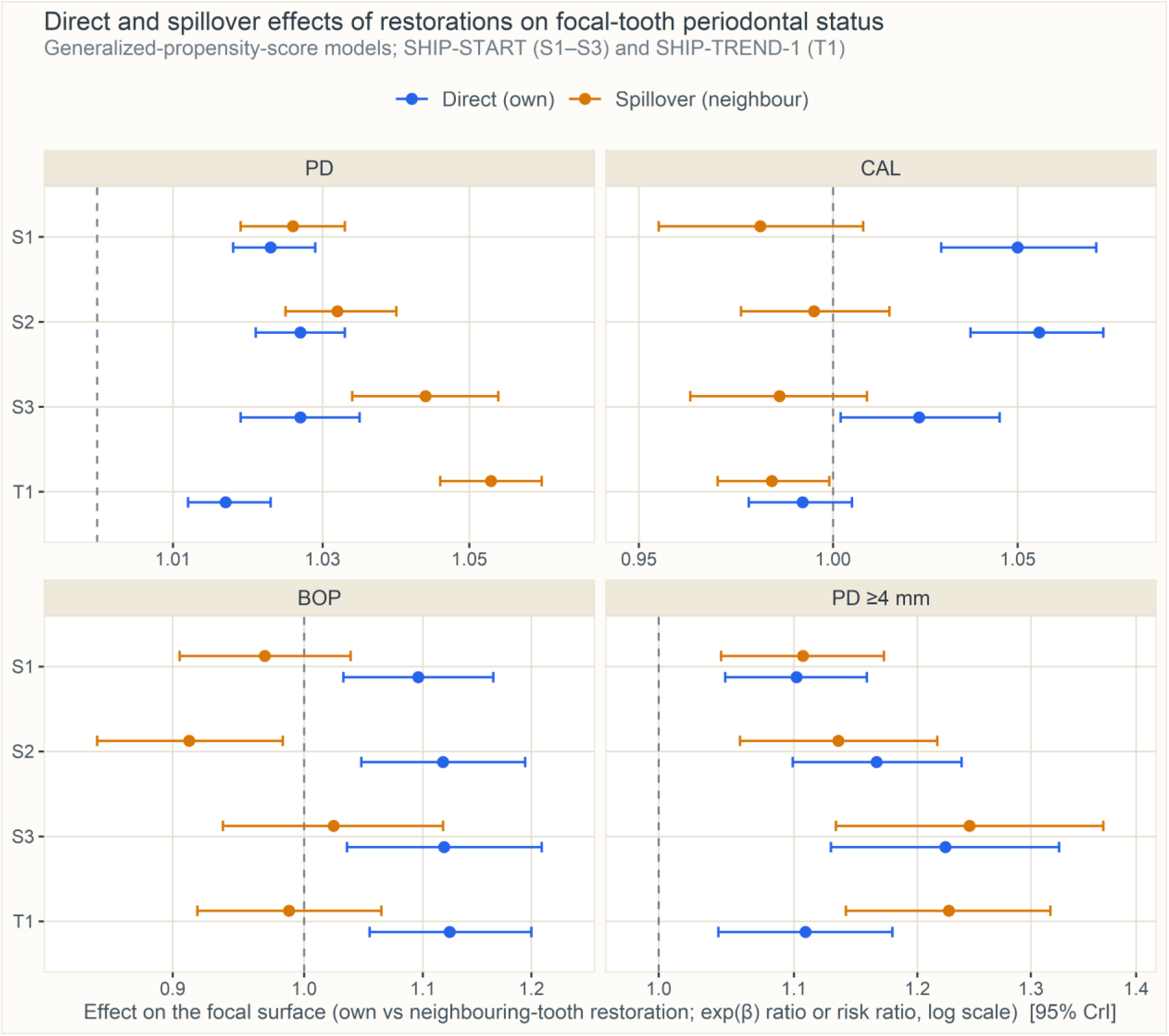
Direct and spillover effects of dental restorations on periodontal status, from the neighbourhood-exposure generalized-propensity-score models. Points are exp(β) (PD, CAL) or RRs (BOP, PD ≥4 mm) with 95% CrI; blue denotes the direct effect of the focal surface’s own restoration and amber the spillover from restorations on neighbouring teeth, across the three SHIP-START follow-ups and the SHIP-TREND replication (log scale). S1–S3 are successive follow-ups of the same cohort; TREND-1 is an independent replication.

Both corroborating estimators reproduced it (Figure 3; Supplementary Table S8). In the joint correlated-random-effects model the indirect effect remained positive (exp(β) 1.009 [1.007, 1.012] at SHIP-START-1 to 1.033 [1.030, 1.035] in SHIP-TREND), while the correlation between the mouth-level random effects of outcome and exposure was small and negative (ρ = −0.003, −0.045, −0.054 and −0.143 at SHIP-START-1, -2, -3 and SHIP-TREND-1), indicating little residual mouth-level confounding. The doubly-robust estimator likewise gave a positive average spillover (+0.045 mm [0.016, 0.073] to +0.144 mm [0.120, 0.168]). Residual Moran’s I along within-person tooth chains was slightly negative in every wave (−0.06 to −0.03) and changed little after neighbourhood adjustment (Supplementary Table S6), arguing against a strong unmodelled contagion process. The three SHIP-START estimates are successive follow-ups of one cohort (1,397 participants appear in all three), not independent replications; SHIP-TREND is the independent replication.

**Figure 3.**
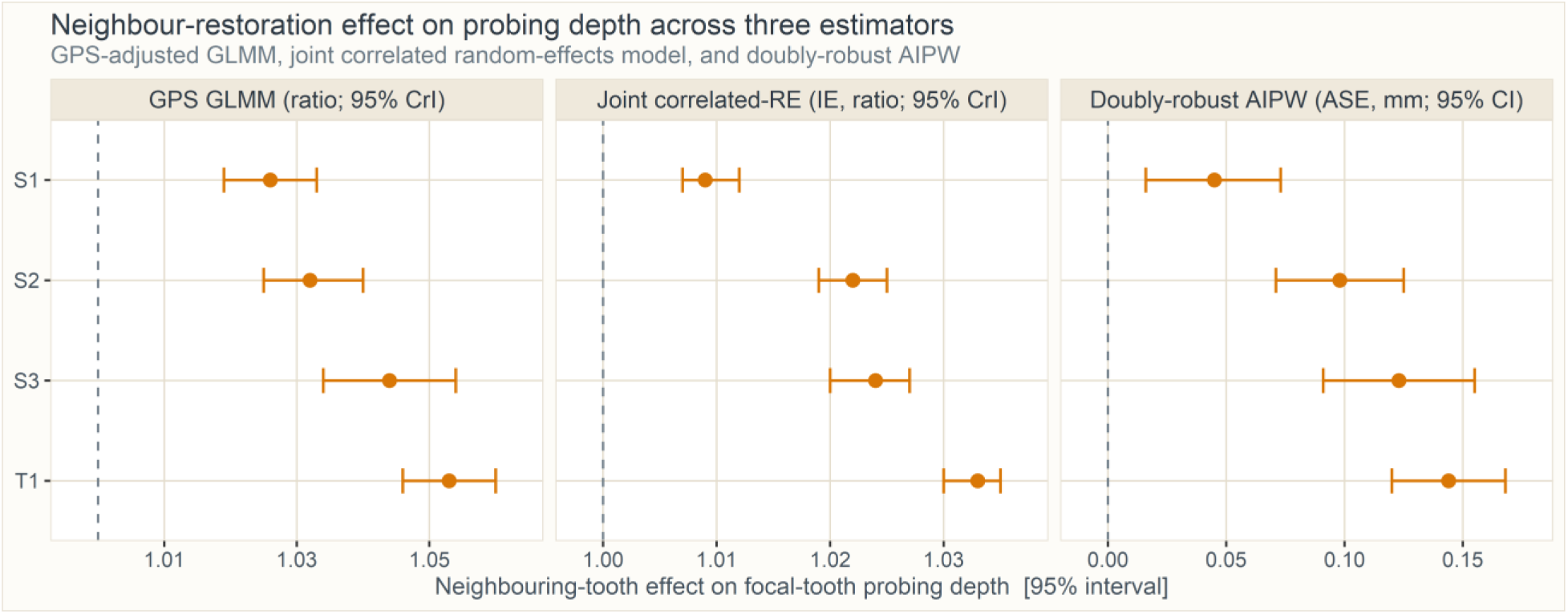
Neighbouring-restoration effect on PD across three estimators: the generalized-propensity-score mixed model (ratio; 95% CrI), the joint correlated-random-effects model (indirect effect; ratio; 95% CrI) and the doubly-robust AIPW estimator (average spillover effect; mm; 95% CI), across SHIP-START and SHIP-TREND. The neighbouring-tooth effect is positive on all three.

### The spillover is concentrated at the neighbouring tooth

Resolving the neighbourhood by tooth-lag, the generalized-propensity-score effect on PD fell from exp(β) 1.025–1.053 at lag 1 to 1.011–1.018 two teeth away and 1.007–1.019 three teeth away, with three of four lag-3 CrIs still excluding 1 and the SHIP-TREND-1 lag-3 estimate exceeding its lag-2 value — attenuation rather than elimination. The deep-pocket RR fell from 1.09–1.20 at lag 1 toward the null, non-monotonically in SHIP-TREND-1 (1.199, 1.073 and 1.128 [1.045, 1.218] at lags 1–3). The mutual GLMM and the joint correlated-random-effects model reached the null by lag 3 (exp(β) 0.98–0.99 and 0.99–1.00), and the doubly-robust spillover declined from +0.06 to +0.15 mm at lag 1 to slightly negative values at lag 3 (−0.06 to −0.03 mm). A CAL spillover at lag 1 appeared only in the mutual GLMM (1.072–1.094) and weakly in the joint correlated-random-effects model (1.006–1.023); it was absent in the generalized-propensity-score model and appeared only inconsistently, in two of four waves, in the doubly-robust estimator (+0.072 mm [0.005, 0.138] at SHIP-START-3 and +0.066 mm [0.022, 0.111] in SHIP-TREND). The spillover was thus concentrated at the neighbouring tooth, reaching the null by lag 3 in the mutual-GLMM and joint correlated-random-effects models while remaining slightly above 1 under the generalized propensity score (Figure 4; Supplementary Figures S1–S2, Table S9); untreated caries showed a concordant decay under the mutual-GLMM, joint correlated-random-effects and doubly-robust estimators, though its imprecise generalized-propensity-score estimates did not decline monotonically (Supplementary Table S10).

**Figure 4.**
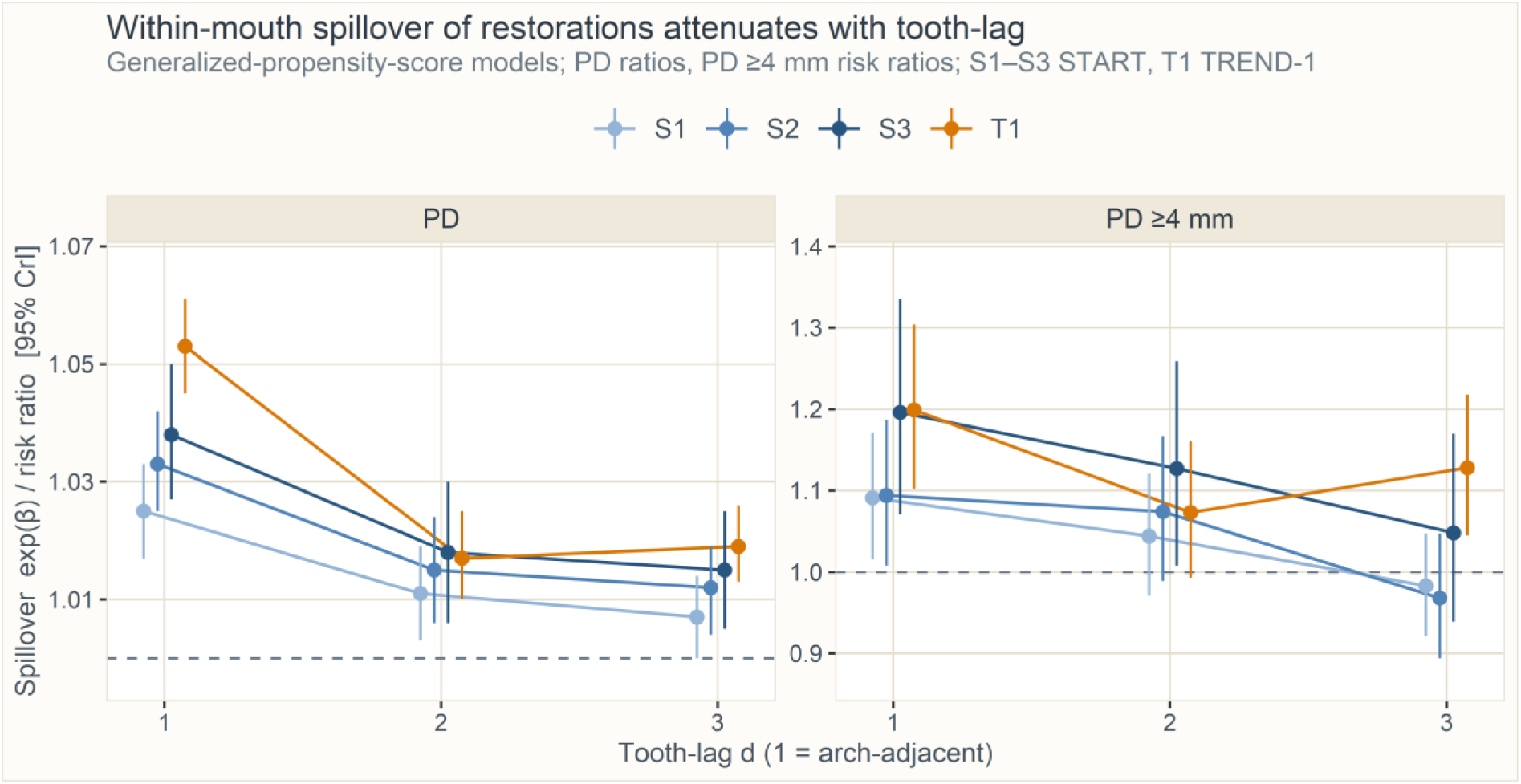
Within-mouth spillover of restorations on PD and deep pockets by tooth-lag. Each panel shows the generalized-propensity-score spillover effect (exp(β) for PD; RR for PD ≥4 mm; 95% CrI) at the neighbouring tooth (lag 1) and at teeth two and three positions away (lags 2–3), across SHIP-START (S1–S3) and SHIP-TREND (T1); S1–S3 are successive follow-ups of the same cohort and TREND-1 is an independent replication. The spillover is concentrated at the immediately neighbouring tooth and attenuates toward the null by lag 2–3. The corresponding CAL and BOP panels appear in Supplementary Figure S2.

### Sensitivity and falsification analyses

Adjusting for tooth type left the primary spillover unchanged (PD exp(β) 1.026–1.050 vs 1.026–1.053) but roughly halved the excess of the mutual-GLMM lag-1 estimates over 1 (SHIP-TREND 1.091 to 1.053) and removed their implausible protective lag-3 values. The spillover was present at both approximal (1.025–1.041) and smooth focal surfaces (1.037– 1.084) and did not sharpen when the exposure was restricted to the facing surface of the neighbouring tooth (0.993–1.015). Restorations placed during follow-up showed no neighbourhood effect (0.98–1.00) while the baseline neighbourhood effect persisted in the same model (1.03–1.05), supporting the baseline, time-integrated exposure as primary. The opposing-tooth negative control was null to slightly protective (PD 0.982–0.986) and left the neighbourhood effect unchanged (Supplementary Section I, Tables S11–S15).

### Confirmatory analysis: untreated caries

Untreated caries, analysed identically, was directionally concordant but imprecise: the PD spillover was positive in the larger waves (neighbouring-tooth exp(β) 1.106 [1.025, 1.194] at SHIP-START-2 and 1.171 [1.052, 1.303] in SHIP-TREND), with a concordant doubly-robust estimate (+0.159 mm [0.034, 0.284] and +0.234 mm [0.108, 0.360]), whereas the own-caries direct effects were unstable (BOP RR 1.221 [0.944, 1.580] at SHIP-START-2 versus 0.772 [0.484, 1.233] in SHIP-TREND). Intervals were wide because untreated caries is rare here (0.1–0.3% of surfaces; Supplementary Table S1) and weighting left residual imbalance (Supplementary Figures S3–S4, Tables S16–S20).

## Discussion

In two population-based cohorts, a focal surface’s own restoration was associated with worse periodontal status at that surface, most strongly for crowns (PD exp(β) up to 1.10), reproducing the established direct association prospectively. The novel finding concerns spillover. Restoration of neighbouring teeth independently raised focal-tooth PD and deep- pocket risk; this persisted after generalized-propensity-score adjustment and was corroborated by the other two estimators. It was specific to PD — CAL and BOP showed no consistent spillover — and untreated caries reproduced it directionally. By tooth-lag it localised to the neighbouring tooth and attenuated by two to three positions, consistent with a spatial-treatment distance decay (Pollmann 2026). Ignoring interference inflated the direct effect only modestly.

Our direct estimates align with recent evidence rather than the larger historical effects: the 26-year Scandinavian study (Schätzle et al. 2001), the Dunedin cohort (Broadbent et al. 2006), the Pelotas 1982 birth cohort (Collares et al. 2018), SHIP-TREND (Nafz et al. 2026) and DMS V (Pitchika et al. 2026) all report site-specific deterioration at restored sites, but modern magnitudes are modest. None estimated spillover. By holding the focal surface’s own restoration fixed and varying the neighbourhood, we provide the first such estimates: the neighbouring-tooth effect on PD survives adjustment for the shared propensity to be restored — a local spillover rather than person-level confounding.

This reframes the literature in two ways. First, own-surface estimates are modestly inflated when interference is ignored: dropping the neighbourhood term raised the direct PD effect by a fifth to a half of its excess over 1, so the no-interference assumption underlying Nafz et al. (2026) and Pitchika et al. (2026) biases the direct association upward without overturning it — standard estimators remain close to a meaningful average effect under limited interference (Sävje et al. 2021). Second, it adds a whole-mouth dimension: a restoration’s periodontal footprint extends to the neighbouring tooth.

The direct association is mechanistically well understood: restoration margins facilitate biofilm accumulation and gingival irritation, driving progression from gingivitis to periodontitis (Lang et al. 2009), while subgingival margins disrupt the supracrestal tissue attachment and alter the microbiome (Gargiulo et al. 1961; Gracis et al. 2023). A shared local environment plausibly underlies the spillover: neighbouring teeth share interproximal contacts, the papilla and a continuous supragingival biofilm (Hagenfeld et al. 2019), and inflammatory and immune markers in crevicular fluid are correlated across sites flanking a shared papilla (Lamster et al. 1991), so a rough or poorly contoured margin may act beyond its own site. The interdental unit is, however, one conduit among others: the spillover was no larger at approximal focal surfaces and did not sharpen at the facing surface of the neighbouring tooth — a segment-level rather than contact-specific process. That the spillover appears in PD — the readout of local swelling and pocketing — but not in CAL, which integrates lifetime history, fits a biofilm-mediated rather than an intrinsic effect (Ercoli et al. 2021).

Two intervals ran in the protective direction (CAL in SHIP-TREND; BOP at SHIP-START-2), compatible with the neighbourhood exposure partly proxying care-seeking: mouths with more restored neighbours receive more regular, mouth-wide dental care; consistent with that channel, the opposing-tooth negative control was itself null to slightly protective.

The spillover is small relative to the measurement: +0.05 to +0.14 mm against a probe graduated in 1-mm increments and non-negligible inter-examiner variability, so it is identifiable only as a mean shift over 17,000–32,000 surfaces per wave. Examiner behaviour and probe angulation are correlated within mouths and cannot be excluded, although an artefact confined to the shared interdental space is unlikely given the spillover at smooth focal surfaces.

Strengths include two independent population-based cohorts; prospective measurement of exposures and covariates at baseline and outcomes at follow-up, with adjustment for the baseline outcome; good covariate balance after inverse-probability-of-treatment weighting; calibrated examiners; and estimation of the spillover with three corroborating estimators and a mutual lag-adjusted mixed model.

Several limitations temper the findings. First, the design is observational; the propensity scores and joint model reduce, but cannot eliminate, confounding. Second, and most important, restoration quality (overhangs, margin location, roughness, material) was not recorded, despite being the strongest known determinant of periodontal harm; the exposure is mere presence. Third, participation was incomplete (baseline response 68.8% in SHIP-START, 50.1% in SHIP-TREND), and because healthier individuals participate, effects are probably underestimated; the spillover was larger at later follow-ups, which may reflect differential attrition and cohort ageing (Supplementary Table S2) as well as cumulative exposure. Fourth, restorations are placed, replaced and lost between the examinations, and this drift affects the neighbourhood exposure more than the focal surface; the incident- restoration models avoid it but make the exposure contemporaneous with the outcome and showed no in-interval neighbourhood effect. Fifth, the protocol was partial-mouth, with bleeding recorded only at index teeth; BOP reflects provoked sulcular bleeding and is suppressed by smoking, an imperfect proxy for local inflammation. Sixth, the interference structure is an assumption and the true geometry of any spillover is unknown. Seventh, CAL was not recorded where the cemento-enamel junction was obscured, so those estimates derive from a selected subset. Eighth, untreated caries was rare, so that analysis had modest power and residual imbalance. Finally, both cohorts are of European ancestry, limiting generalisability.

Clinically, a restoration’s periodontal footprint extends modestly beyond the restored surface to the neighbouring tooth, though the direct effect remains dominant. This reinforces the priorities of precise margin placement and finishing, avoidance of overhangs and rough or subgingival margins, and rigorous supportive periodontal care through close prosthodontic– periodontal collaboration. Future studies should add restoration-quality data and the interproximal microbiome of neighbouring sites. In conclusion, dental restorations exert a local within-mouth spillover on the PD of neighbouring teeth, while the direct association persists, modestly attenuated, once that interference is accounted for.

## Author contributions

Sebastian-Edgar Baumeister: Conceptualization, Methodology, Formal analysis, Data curation, Writing – original draft, Writing – review & editing. Daniel Hagenfeld and Michael Nolde: Conceptualization, Writing – review & editing. Stefanie Samietz, Philipp Kanzow, Henry Völzke, Thomas Kocher and Birte Holtfreter: Investigation, Resources, Writing – review & editing. All authors gave final approval and agree to be accountable for all aspects of the work.

## Ethics statement

SHIP-START and SHIP-TREND were positively evaluated by the ethics committee of the University of Greifswald (SHIP-START-2/SHIP-TREND-0: BB 39/08a; SHIP-TREND-1: BB 174/15) and conducted in accordance with the Declaration of Helsinki. All participants were informed about the study protocol and signed the informed consent and the privacy statement.

## Data availability statement

The data that support the findings of this study are available from Forschungsverbund Community Medicine. Restrictions apply to the availability of these data, which were used under license for this study. Data are available from https://transfer.ship-med.uni-greifswald.de/FAIRequest/login with the permission of Forschungsverbund Community Medicine.

## Use of artificial intelligence

A large-language-model assistant (Claude, Anthropic) was used to support code development and editing (checking grammar and typographical errors).

