## Supplement for "Within-mouth spillover of dental restorations on periodontal status"

### Contents


**A. Examinations, measurements and variables**

**B. Statistical and causal-inference details**

**C. Descriptive statistics and attrition**

**Supplementary Table S1 —** Subject-level characteristics of the baseline examinations and of the analytic sample…

**Supplementary Table S2 —** Baseline characteristics of participants retained in the analytic sample versus those examined…

**D. Covariate balance**

**Supplementary Table S3 —** Baseline covariates by own restoration status (restored vs. sound)…

**Supplementary Table S4 —** Baseline covariates by own restoration status before and after inverse-probability-of-treatment…

**E. Link sensitivity for binary-outcome spillover**

**Supplementary Table S5 —** Binary-outcome spillover across alternative links: logit (odds ratio), linear-probability model…

**F. Identification diagnostics**

**Supplementary Table S6 —** Identification diagnostics for the restoration exposure: positivity (own exposure × any neighbour restored)…

**G. Full estimates behind the main-text figures**

**Supplementary Table S7 —** Primary spillover analysis (Figure 2): neighbourhood-exposure mixed model with individual…

**Supplementary Table S8 —** Neighbouring-tooth restoration effect on PD and CAL across three estimators (Figure 3)…

**H. Spillover by tooth-lag (distance decay)**

**Supplementary Figure S1 —** Tooth-lag decay of the PD spillover across the three estimators…

**Supplementary Figure S2 —** Tooth-lag decay of the CAL and BOP spillover of restorations under the neighbourhood-exposure…

**Supplementary Table S9 —** Within-mouth spillover of restorations on periodontal status by tooth-lag, across all…

**Supplementary Table S10 —** Within-mouth spillover of untreated caries on periodontal status by tooth-lag (confirmatory exposure)…

**I. Additional sensitivity analyses**

**Supplementary Table S11 —** Tooth-type sensitivity: within-mouth spillover of restorations before and after adding tooth…

**Supplementary Table S12 —** Surface-stratum sensitivity: the generalized-propensity-score spillover fitted separately…

**Supplementary Table S13 —** Contact-specific neighbourhood exposure: the neighbourhood exposure sharpened…

**Supplementary Table S14 —** Incident-restoration sensitivity: the own and neighbourhood exposures redefined as restorations…

**Supplementary Table S15 —** Negative-control exposure carried by the opposing (antagonist) tooth — same side, opposite jaw…

**J. Confirmatory analysis: untreated caries**

**Supplementary Table S16 —** Direct effect of untreated caries on the periodontal site at the affected surface…

**Supplementary Table S17 —** Spillover from untreated caries on neighbouring teeth: neighbourhood-exposure mixed model…
**Supplementary Table S18 —** Neighbouring-tooth caries effect on PD and CAL across three estimators…
**Supplementary Table S19 —** Baseline covariates by own caries status (untreated caries vs. sound)…
**Supplementary Table S20 —** Baseline covariates by own caries status before and after inverse-probability-of-treatment…
**Supplementary Figure S3 —** Direct and spillover effects of untreated caries on periodontal status…
**Supplementary Figure S4 —** Neighbouring-tooth caries effect on PD across three estimators…
**References**

### A. Examinations, measurements and variables

Reporting followed the Strengthening the Reporting of Observational Studies in Epidemiology (STROBE) guidelines (von Elm et al., 2014). The Study of Health in Pomerania is a population-based cohort study in north-east Germany (Völzke et al., 2022); examinations were performed prospectively at the baseline (SHIP-START-0, 1997–2001; SHIP-TREND-0, 2008–2012) and at the follow-up examinations (SHIP-START-1/-2/-3 and SHIP-TREND-1) by trained, regularly calibrated dentists. All dental examinations were conducted in an illuminated dental chair with the option of aspiration or an air jet; magnification glasses were not used.

#### Periodontal examination (outcomes)

Probing depth (PD) and clinical attachment level (CAL) were measured with a manual periodontal probe (PCP 11 in SHIP-START; PCP-UNC 15 in SHIP-TREND; Hu-Friedy, Chicago, IL, USA) at four sites per tooth — distobuccal, midbuccal, mesiobuccal and midlingual/midpalatinal — under the half-mouth protocol (one randomly selected side; the same side at baseline and follow-up), excluding third molars. Measurements were rounded to the nearest whole millimetre (0.1–0.4 rounded down, 0.5–0.9 rounded up). PD was the distance between the free gingival margin (FGM) and the pocket base. Where the cemento-enamel junction (CEJ) lay sub-gingivally, CAL was PD minus the FGM–CEJ distance; where recession was present, CAL was measured directly as the CEJ–pocket-base distance. Where the CEJ was indistinct (wedge-shaped defects, fillings, or full/partial crown margins), CAL was not recorded. Bleeding on probing (BOP) was recorded at the same four sites on the first incisor, the canine and the first molar of each probed quadrant; if a reference tooth was missing, the next distally located tooth was assessed. From these, four surface-level outcomes were analysed: PD (mm), CAL (mm), BOP (present/absent) and the presence of a deep pocket (PD ≥4 mm). Periodontal sites were allocated to tooth surfaces by proximity (disto-buccal site → distal surface; mid-buccal → buccal; mesio-buccal → mesial; mid-palatinal/lingual → palatinal/lingual).

#### Caries and restoration assessment (exposures)

Coronal caries and restoration status were assessed visually with a periodontal probe (PCP-UNC 15) touching the surface softly, excluding third molars, at the surface level (distal, buccal, mesial, palatinal/lingual; occlusal surfaces were recorded but not used here, as they do not border the gingival margin), under the same half-mouth protocol. Following Nafz et al. (2026), each surface was classified as sound, primary caries (dentine caries; enamel defects excluded), filled, filled with secondary caries, or missing; information on full or partial crowns (including telescopic crowns and bridges) was taken from the dental-status assessment. From this, two disjoint binary baseline exposures were defined at the focal surface: untreated caries (decayed surfaces — primary caries or filled-with-secondary-caries lesions; caries activity was not recorded) versus sound; and restoration (filled or crowned) versus sound. Restored surfaces were excluded from the caries analysis and carious surfaces from the restoration analysis, so the two exposures do not overlap. The own exposure A is the focal-surface status; the neighbourhood exposure is derived from the neighbouring teeth (below).

#### Examiner calibration and reliability

In each cohort, dental examinations were conducted by six calibrated examiners who, in calibration exercises, repeatedly examined five reference persons not connected to the study. In SHIP-TREND-0, intra-rater correlations were 0.67–0.89 for CAL and 0.68–0.88 for PD (inter-rater 0.70

and 0.72); Cohen's κ was 0.83–1.00 (intra) and 0.72–1.00 (inter) for coronal caries and 0.93–0.99 (intra) and 0.94–0.98 (inter) for tooth status. In SHIP-TREND-1, intra-rater correlations were 0.90–0.96 for CAL and 0.77–0.91 for PD (inter-rater 0.86–0.94 and 0.63–0.85); κ was 0.93–1.00 (intra) and 0.84–0.98 (inter) for caries and 0.97–1.00 (intra) and 0.91–0.96 (inter) for tooth status. SHIP-START examinations followed the same calibrated-examiner protocol.

**Covariates**

Covariates were measured at the baseline examination and harmonised across cohorts. From the computer-assisted personal interview: age (years); sex; school education (<10 / 10 / >10 years); smoking status (never / former / current) and pack-years; toothbrushing frequency (≥2×/day vs. less); daily use of interdental cleaning aids (Holtfreter et al., 2024); and a dental visit within the last 12 months. Body height and weight were measured with calibrated scales and the body mass index computed as weight/height$^2$ (kg/m$^2$). Known diabetes mellitus was defined as a physician's diagnosis or antidiabetic medication (Anatomical Therapeutic Chemical code A10). Probing site was included as a design covariate. Regular professional cleaning and self-reported periodontal treatment were not recorded in SHIP-START-0 and were omitted for cross-cohort comparability.

**Neighbourhood (interference) structure**

For each focal tooth, the interference neighbourhood comprised the neighbouring teeth — the immediately mesial and distal positions within the same quadrant (positions ± 1). The tooth-level degree d is the number of such neighbours present (0, 1 or 2); teeth with degree 0 were excluded from the spillover analyses. The neighbourhood exposure $A_{N(i)}$ is the mean exposure of the neighbouring teeth (a proportion in {0, ½, 1}); the neighbour count G is the number of exposed neighbouring teeth. The neighbourhood exposure is the key interference variable: under no interference its effect on the focal periodontal outcome is zero.

### B. Statistical and causal-inference details

Throughout, $i$ indexes a tooth surface, $j$ a tooth, and $k$ a mouth (the cluster). $A_i \in \{0,1\}$ is the focal-surface (own) exposure, $A_{N(i)}$ the neighbourhood exposure and $G_i$ the number of exposed neighbouring teeth; $Y_i$ is the follow-up periodontal outcome, $X_i$ the harmonised baseline covariate vector (Section A), and $g(\cdot)$ the model link. Under partial interference, interference occurs within but not between mouths. The direct effect is the effect of the focal surface's restoration on the periodontal site at that surface; the spillover (indirect) effect is the effect of restorations on neighbouring teeth on the focal site. The direct effect was estimated by a generalized linear mixed model (GLMM) with random intercepts for person, jaw and tooth (main text); the three estimating strategies below identify the spillover effect under complementary assumptions.

**Primary spillover: generalized propensity-score model (Forastiere et al., 2022)**

Confounding of the two exposures is addressed through two propensity scores. The individual propensity score is the probability of own exposure given covariates,

$$\varphi(X_i) = P(A_i = 1 \mid X_i) = \text{logit}^{-1}(X_i^T \eta), \qquad \text{(S1)}$$

and the neighbourhood generalized propensity score is the Hirano–Imbens conditional density of the observed neighbourhood exposure given own exposure and covariates,

$$\lambda(a \mid A_i, X_i) = \frac{1}{\sqrt{2\pi}\sigma} \exp\left\{\frac{-(a - X_i^T \delta - \zeta A_i)^2}{2\sigma^2}\right\}, \qquad \text{(S2)}$$

evaluated at $a = A_{N(i)}$. The outcome model is a GLMM including own and neighbourhood exposure, natural cubic splines $s_1$, $s_2$ of the two estimated scores, the covariates, the baseline outcome, and mouth/jaw/tooth random intercepts,

$$g(E[Y_i \mid b]) = \alpha + \beta A_i + \gamma A_{N(i)} + s_1(\hat{\varphi}_i) + s_2(\hat{\lambda}_i) + X_i^T \theta + b_k + b_{jk} + b_{ijk}, \quad \text{(S3)}$$

with $g = \log$ for the Gamma outcomes (PD, CAL) and the Poisson working model (BOP, PD ≥4 mm; giving risk ratios, RR). The direct effect is $\exp(\beta)$ and the spillover effect is $\exp(\gamma)$; under no interference $\gamma = 0$.

**Corroboration I: joint correlated random-effects model (McNealis et al., 2026)**

To separate genuine local spillover from shared mouth-level confounding, the outcome and the own exposure are modelled jointly with correlated mouth-level random effects,

$$g\left(E[Y_i \mid b_k^y]\right) = \alpha_y + \beta A_i + \psi G_i + \xi A_i G_i + X_i^T \theta_y + b_k^y, \qquad \text{(S4a)}$$

$$\text{logit} P(A_i = 1 \mid b_k^z) = \alpha_z + X_i^T \theta_z + b_k^z, \text{(S4b)}$$

where the mouth random effects are bivariate normal with an unrestricted covariance,

$$\begin{pmatrix} b_k^y \\ b_k^z \end{pmatrix} \sim N(\mathbf{0}, \Sigma), \Sigma = \begin{pmatrix} \tau_y^2 & \rho \tau_y \tau_z \\ \rho \tau_y \tau_z & \tau_z^2 \end{pmatrix}. \quad \text{(S5)}$$

The correlation $\rho$ (= $\Sigma_{12}$ normalised) captures the shared mouth-level propensity that would otherwise be mistaken for spillover. The indirect (spillover) effect is the contrast of expected focal outcomes under a Bernoulli allocation in which each neighbour is exposed independently with probability $\pi$,

$$\text{IE}(\pi_1, \pi_0) = E_{G \sim \text{Bin}(d_i, \pi_1)}[Y_i] - E_{G \sim \text{Bin}(d_i, \pi_0)}[Y_i], \qquad \text{(S6)}$$

reported at $\pi_1 = 0.7$ versus $\pi_0 = 0.3$ ($d_i$ = degree). Because the linear predictor is linear in the neighbour count, the IE has the closed form $\exp\{\psi \bar{d}(\pi_1 - \pi_0)\}$ on the ratio scale.

**Corroboration II: doubly-robust estimation (Qu et al., 2026)**

As a doubly-robust check, the joint exposure $W_i = (A_i, G_i)$ is modelled with a generalized propensity $\pi(w; X_i) = P(W_i = w \mid X_i)$ (the joint propensity of Qu et al., 2026) and an outcome regression $m(w, X_i) = E[Y_i \mid W_i = w, X_i]$. The augmented inverse-probability-weighted estimator of the mean potential outcome under joint exposure $w$ is

$$\hat{\mu}(w) = \frac{1}{n}\sum_{i=1}^{n}\left[\frac{I(W_i=w)}{\hat{\pi}(w;X_i)}\{Y_i - \hat{m}(w, X_i)\} + \hat{m}(w, X_i)\right], \quad \text{(S7)}$$

and the average spillover effect (Qu's $\tau(0; 1,0)$ ) contrasts an exposed versus unexposed neighbourhood at fixed (unexposed) own status,

$$\widehat{\mathrm{ASE}} = \hat{\mu}(0,1) - \hat{\mu}(0,0). \qquad \text{(S8)}$$

This estimator is consistent if either the propensity model $\hat{\pi}$ or the outcome model $\hat{\mu}$ is correctly specified (double robustness); standard errors use the cluster-robust (mouth-level) influence function.

**Identification and estimation**

Identification rests on partial interference (no between-mouth interference), conditional ignorability of the own and neighbourhood exposures given $X$ (Forastiere et al., 2022), and positivity — each own × any-neighbour-exposed combination is observed across covariate strata (Supplementary Table S6). The joint model additionally separates local spillover from cluster confounding through $\rho$, and the AIPW estimator is valid under single-model misspecification. The two random-effects models (S3 and S4–S5) were fitted by integrated nested Laplace approximation (R-INLA). The PD spillover was cross-validated against exact Markov-chain Monte Carlo (the No-U-Turn sampler in Stan via nutpie), which agreed with INLA to three decimal places (including the random-effect correlation $\rho$); the binary outcomes of the joint model, which did not converge under INLA, were checked by exact MCMC (nutpie) as a convergence check and are not tabulated. Estimates are posterior summaries with 95% credible intervals (CrI); the doubly-robust AIPW estimator is frequentist and is reported with 95% confidence intervals (CI).

### C. Descriptive statistics and attrition

**Supplementary Table S1.** Subject-level characteristics of the baseline examinations and of the analytic sample at each follow-up wave: participants, teeth and surfaces analysed; teeth and surfaces per participant; age, sex, education, smoking, body mass index, diabetes and oral-hygiene/dental-attendance covariates; mean (SD) PD and CAL at baseline and follow-up; prevalence of PD ≥4 mm and of BOP; and the percentage of restored and of untreated-carious surfaces. Values are n, mean (SD) or %.

| Characteristic | START-0 | START-1 | START-2 | START-3 | TREND-0 | TREND-1 |
|---|---|---|---|---|---|---|
| Participants, n | 3,526 | 2,584 | 1,903 | 1,353 | 3,496 | 2,066 |
| Teeth, n | 34,148 | 25,237 | 18,844 | 13,444 | 36,607 | 22,125 |
| Surfaces, n | 118,307 | 87,627 | 65,434 | 46,677 | 145,913 | 88,154 |
| Teeth per participant | 9.7 (3.7) | 9.8 (3.6) | 9.9 (3.5) | 9.9 (3.5) | 10.5 (3.5) | 10.7 (3.2) |
| Surfaces per participant | 33.6 (13.1) | 33.9 (12.8) | 34.4 (12.4) | 34.5 (12.3) | 41.7 (14.0) | 42.7 (12.9) |
| Age at baseline, years | 46.7 (15.1) | 46.0 (14.0) | 44.5 (13.0) | 42.8 (12.3) | 49.6 (14.7) | 48.3 (13.3) |
| Male, % | 48.6 | 47.4 | 47.0 | 46.5 | 48.5 | 48.3 |
| School education <10 years, % | 32.0 | 27.3 | 21.8 | 16.6 | 17.7 | 11.0 |
| School education 10 years, % | 49.4 | 52.6 | 56.5 | 59.8 | 54.1 | 56.0 |
| School education >10 years, % | 18.6 | 20.1 | 21.8 | 23.7 | 28.2 | 33.0 |
| Never smoker, % | 36.0 | 38.0 | 38.9 | 38.8 | 37.4 | 39.4 |
| Former smoker, % | 32.2 | 33.2 | 33.4 | 34.0 | 35.8 | 38.0 |
| Current smoker, % | 31.8 | 28.8 | 27.7 | 27.2 | 26.7 | 22.7 |
| Pack-years (imputed) | 9.7 (13.0) | 8.9 (12.0) | 8.1 (11.3) | 7.2 (10.3) | 10.0 (12.8) | 8.6 (11.5) |
| BMI, kg/m² | 27.0 (4.7) | 26.9 (4.6) | 26.6 (4.5) | 26.3 (4.4) | 27.8 (5.0) | 27.4 (4.6) |
| Diabetes, % | 8.0 | 6.7 | 4.9 | 3.4 | 9.5 | 7.0 |
| Toothbrushing ≥2×/day, % | 83.6 | 85.1 | 85.9 | 86.7 | 85.4 | 87.6 |
| Interdental care, % | 36.6 | 38.9 | 41.0 | 41.4 | 22.6 | 24.1 |
| Dental visit ≤12 months, % | 89.8 | 90.9 | 92.0 | 92.1 | 90.1 | 92.3 |
| PD at baseline, mm | 2.41 (1.12) | 2.35 (1.05) | 2.29 (0.99) | 2.24 (0.96) | 2.46 (1.04) | 2.38 (0.94) |
| PD at follow-up, mm | — | 2.26 (1.10) | 2.60 (1.02) | 2.45 (1.03) | — | 2.32 (0.95) |
| CAL at baseline, mm | 2.03 (1.95) | 1.90 (1.76) | 1.77 (1.65) | 1.64 (1.57) | 2.01 (1.75) | 1.85 (1.55) |
| CAL at follow-up, mm | — | 1.80 (1.90) | 2.43 (1.75) | 2.15 (1.59) | — | 1.99 (1.48) |
| PD ≥4 mm at baseline, % | 10.0 | 8.7 | 7.3 | 6.3 | 11.3 | 9.1 |
| PD ≥4 mm at follow-up, % | — | 12.7 | 11.7 | 8.5 | — | 8.2 |
| BOP at baseline, % | 33.1 | 31.4 | 29.8 | 28.5 | 22.8 | 20.4 |
| BOP at follow-up, % | — | 21.3 | 23.5 | 22.5 | — | 17.4 |
| Restored surfaces at baseline, % | 27.9 | 27.8 | 27.1 | 26.1 | 27.2 | 26.3 |

| | | | | | | |
|---|---|---|---|---|---|---|
| Restored surfaces at follow-up, % | — | 30.9 | 34.6 | 36.0 | — | 30.0 |
| Untreated caries at baseline, % | 0.3 | 0.3 | 0.2 | 0.2 | 0.1 | 0.1 |

*SHIP-START-0 and SHIP-TREND-0 are the baseline examinations, built through the same pipeline (harmonized covariates complete, focal surface classifiable, neighbourhood exposure defined) from the baseline cross-sectional data; rows referring to the follow-up examination are not defined there and are shown as an em dash. Person-level rows are per participant; surface-level rows are per analysed tooth surface. Counts of participants, teeth and surfaces are the analytic sample with the neighbourhood exposure defined; outcome rows are restricted to surfaces on which that outcome was recorded (BOP at index teeth only; CAL where the cemento-enamel junction was identifiable). Abbreviations: PD = probing depth; CAL = clinical attachment level; BOP = bleeding on probing.*

**Supplementary Table S2.** Baseline characteristics of participants retained in the analytic sample versus those examined at baseline but not contributing to the follow-up wave, for each of the three SHIP-START follow-ups and SHIP-TREND-1. Values are mean (SD) or %.

| Wave | Baseline characteristic | Retained | Lost to follow-up |
|---|---|---|---|
| START-1 | Age at baseline, years | 46.0 (14.0) | 51.1 (17.8) |
| START-1 | Male, % | 47.4 | 51.3 |
| START-1 | School education <10 years, % | 27.3 | 49.4 |
| START-1 | School education 10 years, % | 52.6 | 37.6 |
| START-1 | School education >10 years, % | 20.1 | 13.0 |
| START-1 | Never smoker, % | 38.0 | 31.6 |
| START-1 | Former smoker, % | 33.2 | 30.4 |
| START-1 | Current smoker, % | 28.8 | 38.0 |
| START-1 | BMI, kg/m² | 26.9 (4.6) | 27.5 (5.0) |
| START-1 | Diabetes, % | 6.7 | 13.8 |
| START-1 | Toothbrushing ≥2×/day, % | 85.1 | 78.4 |
| START-1 | Dental visit ≤12 months, % | 90.9 | 85.7 |
| START-1 | Teeth examined at baseline, n | 10.9 (3.1) | 8.0 (4.6) |
| START-1 | Restored surfaces at baseline, % | 32.5 (23.9) | 34.5 (31.4) |
| START-1 | PD at baseline, mm | 2.48 (0.63) | 2.79 (0.95) |
| START-1 | Surfaces with PD ≥4 mm at baseline, % | 11.4 (15.2) | 18.5 (22.9) |
| START-2 | Age at baseline, years | 44.5 (13.0) | 50.8 (17.1) |
| START-2 | Male, % | 47.0 | 50.4 |
| START-2 | School education <10 years, % | 21.8 | 46.9 |
| START-2 | School education 10 years, % | 56.5 | 39.1 |
| START-2 | School education >10 years, % | 21.8 | 14.0 |
| START-2 | Never smoker, % | 38.9 | 33.1 |
| START-2 | Former smoker, % | 33.4 | 31.2 |
| START-2 | Current smoker, % | 27.7 | 35.7 |
| START-2 | BMI, kg/m² | 26.6 (4.5) | 27.6 (5.0) |
| START-2 | Diabetes, % | 4.9 | 13.0 |
| START-2 | Toothbrushing ≥2×/day, % | 85.9 | 80.0 |
| START-2 | Dental visit ≤12 months, % | 92.0 | 86.4 |
| START-2 | Teeth examined at baseline, n | 11.3 (2.7) | 8.6 (4.3) |
| START-2 | Restored surfaces at baseline, % | 32.3 (22.9) | 34.0 (29.7) |
| START-2 | PD at baseline, mm | 2.39 (0.55) | 2.76 (0.89) |
| START-2 | Surfaces with PD ≥4 mm at baseline, % | 9.5 (12.9) | 17.9 (21.6) |
| START-3 | Age at baseline, years | 42.8 (12.3) | 50.3 (16.4) |
| START-3 | Male, % | 46.5 | 49.9 |
| START-3 | School education <10 years, % | 16.6 | 44.0 |
| START-3 | School education 10 years, % | 59.8 | 41.3 |
| START-3 | School education >10 years, % | 23.7 | 14.7 |
| START-3 | Never smoker, % | 38.8 | 34.5 |
| START-3 | Former smoker, % | 34.0 | 31.4 |
| START-3 | Current smoker, % | 27.2 | 34.1 |
| START-3 | BMI, kg/m² | 26.3 (4.4) | 27.5 (4.9) |
| START-3 | Diabetes, % | 3.4 | 12.0 |
| START-3 | Toothbrushing ≥2×/day, % | 86.7 | 80.9 |
| START-3 | Dental visit ≤12 months, % | 92.1 | 87.7 |
| START-3 | Teeth examined at baseline, n | 11.6 (2.4) | 9.0 (4.2) |
| START-3 | Restored surfaces at baseline, % | 31.9 (22.5) | 33.8 (28.4) |
| START-3 | PD at baseline, mm | 2.35 (0.52) | 2.70 (0.84) |
| START-3 | Surfaces with PD ≥4 mm at baseline, % | 8.5 (11.7) | 16.5 (20.5) |
| TREND-1 | Age at baseline, years | 48.3 (13.3) | 52.3 (16.4) |

| | | | |
|---|---|---|---|
| TREND-1 | Male, % | 48.3 | 49.4 |
| TREND-1 | School education <10 years, % | 11.0 | 28.4 |
| TREND-1 | School education 10 years, % | 56.0 | 50.7 |
| TREND-1 | School education >10 years, % | 33.0 | 20.8 |
| TREND-1 | Never smoker, % | 39.4 | 34.0 |
| TREND-1 | Former smoker, % | 38.0 | 34.0 |
| TREND-1 | Current smoker, % | 22.7 | 32.0 |
| TREND-1 | BMI, kg/m² | 27.4 (4.6) | 28.4 (5.6) |
| TREND-1 | Diabetes, % | 7.0 | 14.2 |
| TREND-1 | Toothbrushing ≥2×/day, % | 87.6 | 81.7 |
| TREND-1 | Dental visit ≤12 months, % | 92.3 | 86.7 |
| TREND-1 | Teeth examined at baseline, n | 11.8 (2.5) | 9.5 (4.2) |
| TREND-1 | Restored surfaces at baseline, % | 31.8 (22.9) | 36.0 (30.3) |
| TREND-1 | PD at baseline, mm | 2.48 (0.55) | 2.74 (0.85) |
| TREND-1 | Surfaces with PD ≥4 mm at baseline, % | 11.4 (15.6) | 18.5 (23.0) |

*Participants (retained vs lost): START-1: 2584 retained vs 1157 lost; START-2: 1903 retained vs 1838 lost; START-3: 1353 retained vs 2388 lost; TREND-1: 2066 retained vs 1555 lost. Denominators vary slightly by characteristic because of item non-response at baseline. Abbreviations: PD = probing depth.*

### D. Covariate balance

**Supplementary Table S3.** Baseline covariates by own restoration status (restored vs. sound) before and after inverse-probability-of-treatment weighting, SHIP-START. Values are the mean (SD) of each continuous covariate and the percentage in each category of the categorical covariates, given separately for restored and for sound surfaces, together with the standardized mean difference (SMD) between the two groups before and after weighting.

| Variable | Sound | Restored | SMD | Sound (IPTW) | Restored (IPTW) | SMD (IPTW) |
|---|---|---|---|---|---|---|
| Age, yr | 42.6 (13.6) | 46.2 (13.0) | 0.273 | 43.8 (13.8) | 44.2 (12.9) | 0.034 |
| Male, % | 50.7 | 40.7 | -0.203 | 47.8 | 47.7 | -0.002 |
| School <10 yr, % | 21.4 | 24.1 | 0.063 | 22.4 | 23.0 | 0.015 |
| School 10 yr, % | 56.5 | 53.3 | -0.065 | 55.6 | 55.4 | -0.004 |
| School >10 yr, % | 22.0 | 22.6 | 0.014 | 22.1 | 21.6 | -0.011 |
| Never smoker, % | 37.3 | 39.5 | 0.044 | 38.0 | 37.7 | -0.006 |
| Former smoker, % | 32.9 | 32.3 | -0.012 | 32.7 | 33.1 | 0.007 |
| Current smoker, % | 29.8 | 28.2 | -0.035 | 29.3 | 29.2 | -0.002 |
| Pack-years | 8.4 (11.2) | 8.3 (11.2) | -0.003 | 8.4 (11.3) | 8.5 (11.0) | 0.011 |
| BMI, kg/m2 | 26.6 (4.6) | 26.6 (4.5) | 0.002 | 26.6 (4.6) | 26.6 (4.5) | 0.014 |
| Diabetes, % | 5.3 | 5.5 | 0.008 | 5.4 | 5.5 | 0.006 |
| Toothbrushing >=2x/day, % | 84.3 | 88.5 | 0.122 | 85.5 | 85.5 | -0.000 |
| Interdental care, % | 39.4 | 44.3 | 0.100 | 40.8 | 41.1 | 0.005 |
| Dental visit <=12 mo, % | 89.6 | 94.9 | 0.200 | 91.2 | 92.1 | 0.031 |

*Surface-level values; subject-level characteristics in Table S1. Abbreviations: IPTW = inverse-probability-of-treatment weighting; SMD = standardized mean difference.*

**Supplementary Table S4.** Baseline covariates by own restoration status before and after inverse-probability-of-treatment weighting, SHIP-TREND. Values are the mean (SD) of each continuous covariate and the percentage in each category of the categorical covariates, given separately for restored and for sound surfaces, together with the standardized mean difference (SMD) between the two groups before and after weighting.

| Variable | Sound | Restored | SMD | Sound (IPTW) | Restored (IPTW) | SMD (IPTW) |
|---|---|---|---|---|---|---|
| Age, yr | 44.9 (13.2) | 51.0 (12.0) | 0.484 | 46.8 (13.5) | 47.8 (12.0) | 0.081 |
| Male, % | 50.8 | 42.7 | -0.162 | 48.7 | 48.6 | -0.002 |
| School <10 yr, % | 8.8 | 10.6 | 0.058 | 9.5 | 10.1 | 0.020 |
| School 10 yr, % | 55.6 | 57.0 | 0.029 | 56.0 | 56.9 | 0.019 |
| School >10 yr, % | 35.6 | 32.5 | -0.066 | 34.6 | 33.0 | -0.033 |
| Never smoker, % | 39.1 | 41.0 | 0.039 | 39.6 | 39.6 | -0.001 |
| Former smoker, % | 36.8 | 38.8 | 0.041 | 37.4 | 38.0 | 0.011 |
| Current smoker, % | 24.1 | 20.2 | -0.094 | 23.0 | 22.5 | -0.012 |
| Pack-years | 8.1 (10.4) | 8.3 (11.4) | 0.023 | 8.2 (10.8) | 8.4 (11.1) | 0.019 |
| BMI, kg/m2 | 27.0 (4.5) | 27.5 (4.5) | 0.103 | 27.2 (4.5) | 27.3 (4.4) | 0.023 |
| Diabetes, % | 5.7 | 6.7 | 0.042 | 6.0 | 6.2 | 0.011 |
| Toothbrushing >=2x/day, % | 86.9 | 89.1 | 0.068 | 87.4 | 87.4 | -0.001 |
| Interdental care, % | 22.8 | 26.7 | 0.089 | 23.9 | 24.5 | 0.014 |
| Dental visit <=12 mo, % | 91.2 | 94.6 | 0.136 | 92.1 | 92.7 | 0.021 |

*Surface-level values; subject-level characteristics in Table S1. Abbreviations: IPTW = inverse-probability-of-treatment weighting; SMD = standardized mean difference.*

### E. Link sensitivity for binary-outcome spillover

**Supplementary Table S5.** Binary-outcome spillover across alternative links: logit (odds ratio), linear-probability model (risk difference) and probit (coefficient), with 95% CrI, for restoration and caries exposures.

| Exposure | Outcome | Wave | Effect | Logit (OR) | LPM (risk diff.) | Probit (coef.) |
|---|---|---|---|---|---|---|
| Caries | BOP | START-1 | Spillover (neighbour) | 0.965 [0.403, 2.308] | -0.021 [-0.12, 0.078] | -0.012 [-0.517, 0.494] |
| Caries | BOP | START-2 | Spillover (neighbour) | 1.447 [0.54, 3.879] | 0.042 [-0.102, 0.186] | 0.232 [-0.357, 0.822] |
| Caries | BOP | START-3 | Spillover (neighbour) | 3.155 [0.997, 9.984] | 0.195 [0.027, 0.363] | 0.666 [-0.004, 1.335] |
| Caries | BOP | TREND-1 | Spillover (neighbour) | 2.982 [0.603, 14.743] | 0.161 [-0.028, 0.351] | 0.567 [-0.365, 1.5] |
| Caries | BOP | START-1 | Direct (own) | 1.518 [0.791, 2.913] | 0.062 [-0.013, 0.137] | 0.267 [-0.114, 0.648] |
| Caries | BOP | START-2 | Direct (own) | 1.242 [0.605, 2.551] | 0.041 [-0.065, 0.147] | 0.129 [-0.301, 0.558] |
| Caries | BOP | START-3 | Direct (own) | 1.494 [0.614, 3.635] | 0.063 [-0.066, 0.191] | 0.196 [-0.322, 0.714] |
| Caries | BOP | TREND-1 | Direct (own) | 0.11 [0.028, 0.428] | -0.241 [-0.379, -0.102] | -1.197 [-1.95, -0.443] |
| Caries | PD ≥4 mm | START-1 | Spillover (neighbour) | 1.019 [0.487, 2.129] | -0.004 [-0.058, 0.049] | 0.052 [-0.359, 0.463] |
| Caries | PD ≥4 mm | START-2 | Spillover (neighbour) | 1.895 [0.736, 4.883] | 0.038 [-0.031, 0.106] | 0.374 [-0.158, 0.907] |
| Caries | PD ≥4 mm | START-3 | Spillover (neighbour) | 1.204 [0.344, 4.22] | -0.009 [-0.082, 0.064] | 0.144 [-0.536, 0.825] |
| Caries | PD ≥4 mm | TREND-1 | Spillover (neighbour) | 6.303 [1.7, 23.373] | 0.107 [0.029, 0.184] | 0.962 [0.243, 1.68] |
| Caries | PD ≥4 mm | START-1 | Direct (own) | 2.072 [1.205, 3.565] | 0.068 [0.027, 0.108] | 0.386 [0.083, 0.689] |
| Caries | PD ≥4 mm | START-2 | Direct (own) | 0.506 [0.242, 1.054] | -0.026 [-0.078, 0.025] | -0.413 [-0.827, 0] |
| Caries | PD ≥4 mm | START-3 | Direct (own) | 0.563 [0.216, 1.464] | -0.021 [-0.074, 0.033] | -0.269 [-0.785, 0.247] |
| Caries | PD ≥4 mm | TREND-1 | Direct (own) | 0.333 [0.105, 1.06] | 0.035 [-0.028, 0.098] | -0.572 [-1.21, 0.066] |
| Restoration | BOP | START-1 | Spillover (neighbour) | 0.917 [0.833, 1.009] | -0.009 [-0.02, 0.002] | -0.046 [-0.101, 0.009] |
| Restoration | BOP | START-2 | Spillover (neighbour) | 0.849 [0.77, 0.937] | -0.022 [-0.035, -0.008] | -0.092 [-0.149, -0.034] |
| Restoration | BOP | START-3 | Spillover (neighbour) | 1.01 [0.899, 1.134] | 0.004 [-0.013, 0.02] | 0.009 [-0.058, 0.077] |
| Restoration | BOP | TREND-1 | Spillover (neighbour) | 0.979 [0.893, 1.073] | -0.001 [-0.012, 0.009] | -0.014 [-0.065, 0.038] |
| Restoration | BOP | START-1 | Direct (own) | 1.166 [1.074, 1.267] | 0.016 [0.006, 0.025] | 0.084 [0.037, 0.132] |
| Restoration | BOP | START-2 | Direct (own) | 1.188 [1.09, 1.295] | 0.022 [0.01, 0.034] | 0.099 [0.049, 0.149] |
| Restoration | BOP | START-3 | Direct (own) | 1.19 [1.075, 1.316] | 0.025 [0.01, 0.039] | 0.098 [0.039, 0.157] |
| Restoration | BOP | TREND-1 | Direct (own) | 1.167 [1.077, 1.263] | 0.018 [0.008, 0.028] | 0.088 [0.042, 0.133] |
| Restoration | PD ≥4 mm | START-1 | Spillover (neighbour) | 1.18 [1.087, 1.28] | 0.009 [0.003, 0.015] | 0.092 [0.047, 0.137] |
| Restoration | PD ≥4 mm | START-2 | Spillover (neighbour) | 1.248 [1.137, 1.371] | 0.014 [0.008, 0.021] | 0.118 [0.067, 0.17] |
| Restoration | PD ≥4 mm | START-3 | Spillover (neighbour) | 1.369 [1.214, 1.544] | 0.017 [0.009, 0.024] | 0.168 [0.104, 0.233] |
| Restoration | PD ≥4 mm | TREND-1 | Spillover (neighbour) | 1.365 [1.24, 1.501] | 0.015 [0.01, 0.02] | 0.167 [0.116, 0.218] |
| Restoration | PD ≥4 mm | START-1 | Direct (own) | 1.187 [1.108, 1.272] | 0.011 [0.006, 0.016] | 0.088 [0.05, 0.126] |

| | | | | | | |
|---|---|---|---|---|---|---|
| Restoration | PD ≥4 mm | START-2 | Direct (own) | 1.28 [1.184, 1.384] | 0.016 [0.011, 0.022] | 0.141 [0.098, 0.185] |
| Restoration | PD ≥4 mm | START-3 | Direct (own) | 1.342 [1.214, 1.483] | 0.017 [0.011, 0.023] | 0.155 [0.1, 0.209] |
| Restoration | PD ≥4 mm | TREND-1 | Direct (own) | 1.169 [1.08, 1.266] | 0.007 [0.003, 0.011] | 0.08 [0.037, 0.123] |

*Abbreviations: PD = probing depth; BOP = bleeding on probing; CrI = credible interval.*

### F. Identification diagnostics

**Supplementary Table S6.** Identification diagnostics for the restoration exposure: positivity (own exposure × any neighbour restored), mean neighbourhood exposure, its correlation with whole-mouth restoration burden, intraclass correlation (ICC) of neighbourhood exposure, and Moran's I before and after neighbourhood adjustment.

| Wave | Positivity: (0,0) | Positivity: (0,1) | Positivity: (1,0) | Positivity: (1,1) | nb mean (SD) | Corr(nb, mouth%) | ICC of nb | Moran I (before nb adj) | Moran I (after nb adj) | Moran reduction % |
|---|---|---|---|---|---|---|---|---|---|---|
| START-1 | 11,677 (36.0) | 10,208 (31.5) | 1,898 (5.9) | 8,609 (26.6) | 0.439 (0.442) | 0.497 | 0.399 | -0.0571 | -0.0566 | 0.9 |
| START-2 | 8,807 (36.4) | 7,653 (31.6) | 1,452 (6.0) | 6,294 (26.0) | 0.431 (0.441) | 0.493 | 0.382 | -0.0457 | -0.0465 | -1.7 |
| START-3 | 6,429 (37.3) | 5,430 (31.5) | 1,016 (5.9) | 4,372 (25.3) | 0.422 (0.440) | 0.502 | 0.394 | -0.0455 | -0.0444 | 2.3 |
| TREND-1 | 9,857 (34.7) | 9,034 (31.8) | 1,517 (5.3) | 7,999 (28.2) | 0.432 (0.440) | 0.483 | 0.344 | -0.0298 | -0.0289 | 3.1 |

*Positivity cells give the number of surfaces with row percentages in parentheses, i.e. the percentage of that wave's analysed surfaces falling in each own × neighbourhood exposure combination (0 = unexposed, 1 = exposed; the four cells sum to 100% within a wave). Abbreviations: Nb = neighbourhood.*

### G. Full estimates behind the main-text figures

*Figures 2 and 3 of the main text present the primary within-mouth spillover results graphically; the corresponding numeric estimates are tabulated here.*

**Supplementary Table S7.** Primary spillover analysis (Figure 2): neighbourhood-exposure mixed model with individual and neighbourhood generalized-propensity-score adjustment. Direct (own) and spillover (neighbouring-tooth) effects; exp(β) for PD/CAL, RRs for BOP/PD ≥4 mm, with 95% CrI.

| Outcome | Scale | Effect | START-1 | START-2 | START-3 | TREND-1 |
|---|---|---|---|---|---|---|
| BOP | RR | Direct (own) | 1.096 [1.032, 1.164] | 1.118 [1.047, 1.194] | 1.119 [1.035, 1.210] | 1.124 [1.054, 1.200] |
| BOP | RR | Spillover (neighbour) | 0.969 [0.905, 1.038] | 0.912 [0.847, 0.983] | 1.024 [0.937, 1.118] | 0.988 [0.918, 1.064] |
| CAL | ratio | Direct (own) | 1.050 [1.029, 1.072] | 1.056 [1.037, 1.074] | 1.023 [1.002, 1.045] | 0.992 [0.978, 1.005] |
| CAL | ratio | Spillover (neighbour) | 0.981 [0.955, 1.008] | 0.995 [0.976, 1.015] | 0.986 [0.963, 1.009] | 0.984 [0.970, 0.999] |
| PD | ratio | Direct (own) | 1.023 [1.018, 1.029] | 1.027 [1.021, 1.033] | 1.027 [1.019, 1.035] | 1.017 [1.012, 1.023] |
| PD | ratio | Spillover (neighbour) | 1.026 [1.019, 1.033] | 1.032 [1.025, 1.040] | 1.044 [1.034, 1.054] | 1.053 [1.046, 1.060] |
| PD ≥4 mm | RR | Direct (own) | 1.102 [1.048, 1.158] | 1.166 [1.099, 1.238] | 1.224 [1.129, 1.326] | 1.109 [1.043, 1.179] |
| PD ≥4 mm | RR | Spillover (neighbour) | 1.107 [1.045, 1.172] | 1.135 [1.059, 1.217] | 1.245 [1.133, 1.368] | 1.227 [1.141, 1.318] |

*Abbreviations: PD = probing depth; CAL = clinical attachment level; BOP = bleeding on probing; CrI = credible interval; RR = risk ratio.*

**Supplementary Table S8.** Neighbouring-tooth restoration effect on PD and CAL across three estimators (Figure 3): generalized-propensity-score mixed model; joint correlated-random-effects model (indirect effect); doubly-robust AIPW (average spillover effect). 95% CrI for the two Bayesian models; 95% CI for AIPW.

| Outcome | Estimator | START-1 | START-2 | START-3 | TREND-1 |
|---|---|---|---|---|---|
| CAL | Doubly-robust AIPW (ASE, mm) | -0.061 [-0.128, 0.006] | -0.015 [-0.083, 0.054] | 0.055 [-0.020, 0.131] | 0.050 [-0.002, 0.102] |
| CAL | GPS GLMM (neighbour, ratio) | 0.981 [0.955, 1.008] | 0.995 [0.976, 1.015] | 0.986 [0.963, 1.009] | 0.984 [0.970, 0.999] |
| CAL | Joint correlated-RE (IE, ratio) | 1.007 [0.998, 1.016] | 1.034 [1.025, 1.042] | | 1.027 [1.021, 1.034] |
| PD | Doubly-robust AIPW (ASE, mm) | 0.045 [0.016, 0.073] | 0.098 [0.071, 0.125] | 0.123 [0.091, 0.155] | 0.144 [0.120, 0.168] |
| PD | GPS GLMM (neighbour, ratio) | 1.026 [1.019, 1.033] | 1.032 [1.025, 1.040] | 1.044 [1.034, 1.054] | 1.053 [1.046, 1.060] |
| PD | Joint correlated-RE (IE, ratio) | 1.009 [1.007, 1.012] | 1.022 [1.019, 1.025] | 1.024 [1.020, 1.027] | 1.033 [1.030, 1.035] |

*Abbreviations: PD = probing depth; CAL = clinical attachment level; CrI = credible interval; CI = confidence interval; GPS = generalized propensity score; RE = random effects; AIPW = augmented inverse-probability weighting.*

### H. Spillover by tooth-lag (distance decay)

To characterise how far the within-mouth spillover extends along the arch, the neighbourhood exposure was redefined by topological tooth-lag d (d = 1 the neighbouring tooth; d = 2, 3 for teeth two and three positions away within the quadrant; every tooth retains at least one neighbour at each lag, so positivity holds) and each spillover estimator refitted with the lag-d exposure, the mixed and propensity-score models adjusting for lags 1–3 simultaneously so that each lag is estimated holding the others fixed. This adapts the spatial-treatment distance-decay framework of Pollmann (2026) to the discrete dentition and is a descriptive distance-decay extension, not a design-based estimate. Across all estimators and both exposures the spillover is concentrated at the immediately neighbouring tooth and attenuates towards the null by tooth-lag 2–3 (Supplementary Figures S1–S2).

**Supplementary Figure S1.** Tooth-lag decay of the PD spillover across the three estimators (generalized-propensity-score mixed model and joint correlated-random-effects model on the ratio scale; doubly-robust AIPW on the additive millimetre scale), for the restoration and untreated-caries exposures, across SHIP-START (S1–S3) and SHIP-TREND (T1).

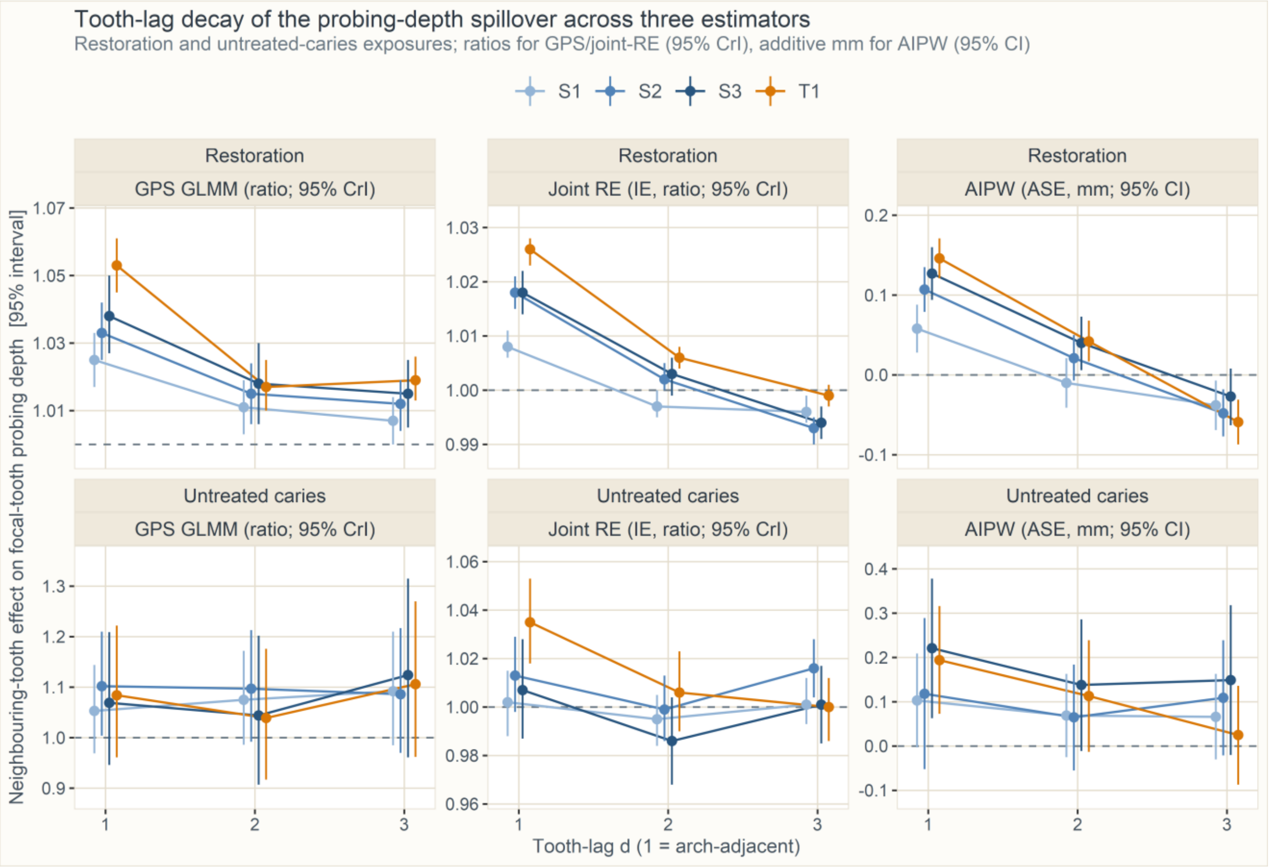

**Supplementary Figure S2.** Tooth-lag decay of the CAL and BOP spillover of restorations under the neighbourhood-exposure generalized-propensity-score model: exp(β) for CAL and RRs for BOP, with 95% CrI, at tooth-lags 1–3 across SHIP-START (S1–S3) and SHIP-TREND (T1). Panels moved from main-text Figure 4, which now shows PD and PD ≥4 mm only; the corresponding joint correlated-random-effects and doubly-robust AIPW estimates are tabulated in Supplementary Table S9.

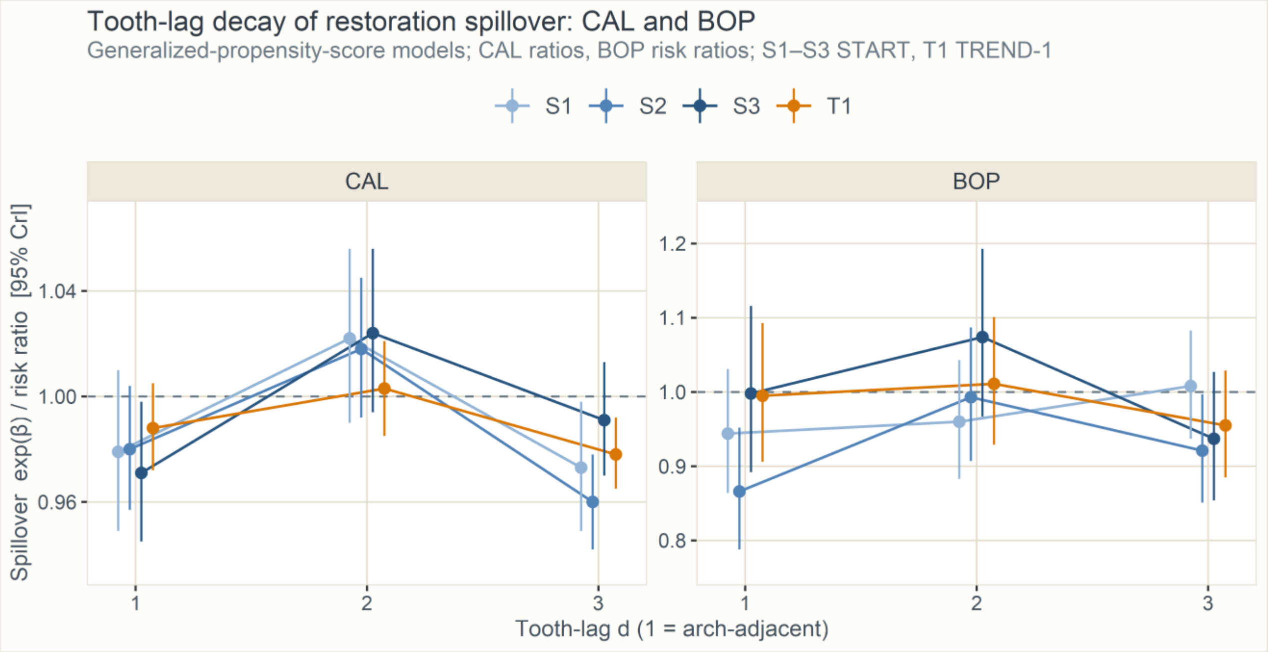

**Supplementary Table S9.** Within-mouth spillover of restorations on periodontal status by tooth-lag, across all estimators (mutual GLMM; Forastiere generalized propensity score; McNealis joint correlated-random-effects model, PD/CAL only; Qu doubly-robust AIPW). Ratios for the mixed/propensity-score and joint-RE models, additive millimetres (PD/CAL) or risk differences (BOP/PD ≥4 mm) for AIPW; 95% CrI for the Bayesian models and 95% CI for AIPW; lags 1–3 mutually adjusted where applicable.

| Outcome | Estimator | Tooth-lag | START-1 | START-2 | START-3 | TREND-1 |
|---|---|---|---|---|---|---|
| BOP | Mutual GLMM | lag 1 | 1.012 [0.932, 1.099] | 0.887 [0.812, 0.970] | 1.078 [0.971, 1.196] | 1.092 [1.001, 1.191] |
| BOP | Mutual GLMM | lag 2 | 1.013 [0.937, 1.096] | 1.037 [0.953, 1.128] | 1.155 [1.048, 1.274] | 1.036 [0.954, 1.124] |
| BOP | Mutual GLMM | lag 3 | 0.874 [0.820, 0.932] | 0.836 [0.781, 0.895] | 0.806 [0.744, 0.874] | 0.775 [0.726, 0.828] |
| BOP | Forastiere GPS | lag 1 | 0.944 [0.864, 1.031] | 0.866 [0.788, 0.952] | 0.998 [0.892, 1.116] | 0.995 [0.906, 1.093] |
| BOP | Forastiere GPS | lag 2 | 0.960 [0.883, 1.043] | 0.993 [0.907, 1.087] | 1.074 [0.967, 1.193] | 1.011 [0.929, 1.101] |
| BOP | Forastiere GPS | lag 3 | 1.008 [0.937, 1.083] | 0.921 [0.851, 0.997] | 0.937 [0.854, 1.027] | 0.955 [0.885, 1.029] |
| BOP | Qu AIPW | lag 1 | 0.011 [-0.005, 0.027] | -0.019 [-0.037, -0.001] | 0.026 [0.005, 0.046] | 0.014 [0.001, 0.026] |
| BOP | Qu AIPW | lag 2 | -0.017 [-0.033, -0.001] | -0.015 [-0.033, 0.003] | 0.021 [0.001, 0.041] | -0.008 [-0.020, 0.005] |
| BOP | Qu AIPW | lag 3 | -0.024 [-0.039, -0.010] | -0.036 [-0.053, -0.018] | -0.029 [-0.048, -0.010] | -0.032 [-0.045, -0.018] |
| CAL | Mutual GLMM | lag 1 | 1.089 [1.060, 1.120] | 1.094 [1.072, 1.117] | 1.090 [1.064, 1.115] | 1.072 [1.056, 1.087] |
| CAL | Mutual GLMM | lag 2 | 1.037 [1.010, 1.065] | 1.038 [1.018, 1.060] | 1.044 [1.021, 1.068] | 1.028 [1.013, 1.042] |
| CAL | Mutual GLMM | lag 3 | 0.931 [0.909, 0.953] | 0.902 [0.886, 0.919] | 0.934 [0.916, 0.953] | 0.928 [0.916, 0.940] |
| CAL | Forastiere GPS | lag 1 | 0.979 [0.949, 1.010] | 0.980 [0.957, 1.004] | 0.971 [0.945, 0.998] | 0.988 [0.972, 1.005] |
| CAL | Forastiere GPS | lag 2 | 1.022 [0.990, 1.056] | 1.018 [0.992, 1.045] | 1.024 [0.994, 1.056] | 1.003 [0.985, 1.021] |
| CAL | Forastiere GPS | lag 3 | 0.973 [0.949, 0.998] | 0.960 [0.942, 0.978] | 0.991 [0.970, 1.013] | 0.978 [0.965, 0.992] |
| CAL | McNealis joint-RE | lag 1 | 1.006 [0.999, 1.019] | 1.023 [1.014, 1.032] | 1.018 [1.006, 1.029] | 1.021 [1.013, 1.027] |
| CAL | McNealis joint-RE | lag 2 | 0.990 [0.981, 0.997] | 0.995 [0.986, 1.003] | 1.008 [0.999, 1.019] | 1.006 [1.000, 1.012] |
| CAL | McNealis joint-RE | lag 3 | 0.971 [0.962, 0.979] | 0.951 [0.944, 0.959] | 0.966 [0.958, 0.974] | 0.963 [0.957, 0.969] |
| CAL | Qu AIPW | lag 1 | -0.038 [-0.097, 0.021] | -0.008 [-0.068, 0.053] | 0.072 [0.005, 0.138] | 0.066 [0.022, 0.111] |
| CAL | Qu AIPW | lag 2 | -0.057 [-0.119, 0.005] | -0.075 [-0.139, -0.011] | 0.005 [-0.062, 0.072] | -0.001 [-0.047, 0.046] |
| CAL | Qu AIPW | lag 3 | -0.132 [-0.195, -0.068] | -0.192 [-0.256, -0.127] | -0.118 [-0.185, -0.051] | -0.137 [-0.187, -0.087] |
| PD | Mutual GLMM | lag 1 | 1.052 [1.044, 1.059] | 1.079 [1.071, 1.088] | 1.081 [1.069, 1.092] | 1.091 [1.083, 1.098] |
| PD | Mutual GLMM | lag 2 | 1.017 [1.010, 1.025] | 1.031 [1.022, 1.039] | 1.033 [1.022, 1.044] | 1.029 [1.022, 1.036] |
| PD | Mutual GLMM | lag 3 | 0.994 [0.988, 1.000] | 0.980 [0.974, 0.987] | 0.987 [0.978, 0.996] | 0.993 [0.987, 0.999] |
| PD | Forastiere GPS | lag 1 | 1.025 [1.017, 1.033] | 1.033 [1.025, 1.042] | 1.038 [1.027, 1.050] | 1.053 [1.045, 1.061] |
| PD | Forastiere GPS | lag 2 | 1.011 [1.003, 1.019] | 1.015 [1.006, 1.024] | 1.018 [1.006, 1.030] | 1.017 [1.010, 1.025] |
| PD | Forastiere GPS | lag 3 | 1.007 [1.000, 1.014] | 1.012 [1.004, 1.019] | 1.015 [1.005, 1.025] | 1.019 [1.013, 1.026] |
| PD | McNealis joint-RE | lag 1 | 1.008 [1.006, 1.011] | 1.018 [1.015, 1.021] | 1.018 [1.014, 1.022] | 1.026 [1.023, 1.028] |
| PD | McNealis joint-RE | lag 2 | 0.997 [0.995, 1.000] | 1.002 [1.000, 1.005] | 1.003 [0.999, 1.006] | 1.006 [1.004, 1.008] |

| | | | | | | |
|---|---|---|---|---|---|---|
| PD | McNealis joint-RE | lag 3 | 0.996 [0.994, 0.999] | 0.993 [0.990, 0.995] | 0.994 [0.991, 0.997] | 0.999 [0.997, 1.001] |
| PD | Qu AIPW | lag 1 | 0.058 [0.028, 0.088] | 0.107 [0.079, 0.135] | 0.127 [0.094, 0.160] | 0.146 [0.121, 0.171] |
| PD | Qu AIPW | lag 2 | -0.010 [-0.041, 0.021] | 0.021 [-0.007, 0.050] | 0.040 [0.006, 0.073] | 0.042 [0.017, 0.068] |
| PD | Qu AIPW | lag 3 | -0.038 [-0.069, -0.007] | -0.048 [-0.077, -0.018] | -0.027 [-0.063, 0.008] | -0.059 [-0.087, -0.031] |
| PD ≥4 mm | Mutual GLMM | lag 1 | 1.217 [1.139, 1.301] | 1.330 [1.231, 1.438] | 1.573 [1.413, 1.752] | 1.506 [1.391, 1.630] |
| PD ≥4 mm | Mutual GLMM | lag 2 | 1.068 [1.002, 1.139] | 1.183 [1.097, 1.276] | 1.250 [1.129, 1.386] | 1.132 [1.049, 1.221] |
| PD ≥4 mm | Mutual GLMM | lag 3 | 0.950 [0.898, 1.006] | 0.840 [0.785, 0.899] | 0.886 [0.808, 0.971] | 0.968 [0.904, 1.036] |
| PD ≥4 mm | Forastiere GPS | lag 1 | 1.091 [1.016, 1.171] | 1.094 [1.008, 1.187] | 1.196 [1.071, 1.335] | 1.199 [1.102, 1.304] |
| PD ≥4 mm | Forastiere GPS | lag 2 | 1.044 [0.971, 1.121] | 1.074 [0.989, 1.167] | 1.127 [1.008, 1.259] | 1.073 [0.993, 1.161] |
| PD ≥4 mm | Forastiere GPS | lag 3 | 0.983 [0.922, 1.047] | 0.968 [0.894, 1.047] | 1.048 [0.939, 1.170] | 1.128 [1.045, 1.218] |
| PD ≥4 mm | Qu AIPW | lag 1 | 0.005 [-0.003, 0.013] | 0.011 [0.003, 0.019] | 0.020 [0.011, 0.029] | 0.020 [0.013, 0.027] |
| PD ≥4 mm | Qu AIPW | lag 2 | -0.010 [-0.018, -0.002] | -0.004 [-0.013, 0.004] | 0.006 [-0.003, 0.015] | 0.002 [-0.005, 0.009] |
| PD ≥4 mm | Qu AIPW | lag 3 | -0.011 [-0.019, -0.002] | -0.016 [-0.025, -0.007] | -0.010 [-0.019, 0.000] | -0.013 [-0.021, -0.006] |

*Abbreviations: PD = probing depth; CAL = clinical attachment level; BOP = bleeding on probing; CrI = credible interval; CI = confidence interval; GPS = generalized propensity score; RE = random effects; AIPW = augmented inverse-probability weighting.*

**Supplementary Table S10.** Within-mouth spillover of untreated caries on periodontal status by tooth-lag (confirmatory exposure), across all estimators; conventions as in Supplementary Table S9.

| Outcome | Estimator | Tooth-lag | START-1 | START-2 | START-3 | TREND-1 |
|---|---|---|---|---|---|---|
| BOP | Mutual GLMM | lag 1 | 1.161 [0.815, 1.656] | 1.036 [0.699, 1.536] | 1.858 [1.218, 2.834] | 1.174 [0.713, 1.933] |
| BOP | Mutual GLMM | lag 2 | 0.798 [0.552, 1.153] | 1.460 [0.989, 2.156] | 0.807 [0.458, 1.424] | 1.364 [0.837, 2.223] |
| BOP | Mutual GLMM | lag 3 | 0.940 [0.728, 1.214] | 1.279 [0.975, 1.678] | 1.285 [0.922, 1.792] | 0.742 [0.477, 1.152] |
| BOP | Forastiere GPS | lag 1 | 0.895 [0.729, 1.100] | 1.881 [0.668, 5.295] | 1.111 [0.849, 1.456] | 1.006 [0.747, 1.354] |
| BOP | Forastiere GPS | lag 2 | 0.731 [0.599, 0.893] | 2.100 [0.622, 7.090] | 0.761 [0.544, 1.064] | 0.881 [0.663, 1.170] |
| BOP | Forastiere GPS | lag 3 | 0.816 [0.704, 0.946] | 0.290 [0.004, 19.709] | 0.981 [0.788, 1.220] | 0.784 [0.632, 0.973] |
| BOP | Qu AIPW | lag 1 | 0.061 [-0.020, 0.141] | 0.033 [-0.038, 0.104] | 0.137 [0.008, 0.265] | 0.054 [-0.028, 0.136] |
| BOP | Qu AIPW | lag 2 | -0.041 [-0.092, 0.010] | 0.007 [-0.086, 0.099] | -0.036 [-0.120, 0.047] | 0.056 [-0.002, 0.114] |
| BOP | Qu AIPW | lag 3 | 0.021 [-0.052, 0.094] | 0.065 [-0.009, 0.138] | 0.132 [0.002, 0.262] | -0.009 [-0.082, 0.064] |
| CAL | Mutual GLMM | lag 1 | 1.030 [0.913, 1.161] | 1.124 [1.021, 1.238] | 1.034 [0.925, 1.157] | 1.089 [0.996, 1.189] |
| CAL | Mutual GLMM | lag 2 | 1.024 [0.915, 1.147] | 1.098 [1.001, 1.204] | 1.061 [0.954, 1.180] | 0.972 [0.890, 1.063] |
| CAL | Mutual GLMM | lag 3 | 0.970 [0.882, 1.066] | 1.082 [1.003, 1.167] | 1.090 [0.999, 1.190] | 0.966 [0.896, 1.042] |
| CAL | Forastiere GPS | lag 1 | 1.048 [0.779, 1.411] | 1.393 [1.090, 1.779] | 1.142 [0.871, 1.498] | 1.244 [0.978, 1.582] |
| CAL | Forastiere GPS | lag 2 | 1.093 [0.806, 1.483] | 1.137 [0.885, 1.460] | 1.520 [1.124, 2.056] | 0.878 [0.685, 1.125] |
| CAL | Forastiere GPS | lag 3 | 1.048 [0.729, 1.508] | 1.180 [0.891, 1.562] | 1.472 [1.043, 2.079] | 1.011 [0.771, 1.326] |
| CAL | McNealis joint-RE | lag 1 | 0.973 [0.929, 1.017] | 1.013 [0.967, 1.063] | 0.990 [0.932, 1.048] | 1.027 [0.985, 1.071] |
| CAL | McNealis joint-RE | lag 2 | 0.987 [0.945, 1.023] | 1.017 [0.978, 1.060] | 0.982 [0.937, 1.028] | 0.985 [0.951, 1.023] |
| CAL | McNealis joint-RE | lag 3 | 0.969 [0.938, 1.004] | 1.021 [0.986, 1.055] | 1.016 [0.971, 1.060] | 0.973 [0.939, 1.006] |
| CAL | Qu AIPW | lag 1 | 0.196 [-0.072, 0.464] | 0.154 [-0.163, 0.471] | 0.189 [-0.291, 0.670] | 0.119 [-0.130, 0.368] |
| CAL | Qu AIPW | lag 2 | 0.151 [-0.047, 0.349] | 0.166 [-0.081, 0.414] | 0.142 [-0.172, 0.456] | 0.067 [-0.186, 0.319] |
| CAL | Qu AIPW | lag 3 | 0.172 [-0.055, 0.399] | 0.245 [-0.044, 0.534] | 0.349 [-0.049, 0.746] | -0.036 [-0.268, 0.195] |
| PD | Mutual GLMM | lag 1 | 1.022 [0.988, 1.057] | 1.048 [1.008, 1.090] | 1.050 [0.995, 1.108] | 1.089 [1.041, 1.140] |
| PD | Mutual GLMM | lag 2 | 1.009 [0.977, 1.042] | 1.033 [0.994, 1.073] | 1.000 [0.949, 1.054] | 1.022 [0.977, 1.070] |
| PD | Mutual GLMM | lag 3 | 1.007 [0.980, 1.035] | 1.042 [1.010, 1.076] | 1.030 [0.986, 1.075] | 1.010 [0.971, 1.050] |
| PD | Forastiere GPS | lag 1 | 1.053 [0.969, 1.144] | 1.102 [1.004, 1.210] | 1.069 [0.946, 1.209] | 1.084 [0.961, 1.222] |
| PD | Forastiere GPS | lag 2 | 1.075 [0.986, 1.172] | 1.097 [0.992, 1.213] | 1.044 [0.907, 1.202] | 1.039 [0.917, 1.176] |
| PD | Forastiere GPS | lag 3 | 1.092 [0.985, 1.210] | 1.086 [0.970, 1.217] | 1.124 [0.961, 1.315] | 1.106 [0.962, 1.270] |
| PD | McNealis joint-RE | lag 1 | 1.002 [0.988, 1.015] | 1.013 [0.998, 1.029] | 1.007 [0.987, 1.028] | 1.035 [1.018, 1.053] |
| PD | McNealis joint-RE | lag 2 | 0.995 [0.984, 1.005] | 0.999 [0.986, 1.013] | 0.986 [0.968, 1.004] | 1.006 [0.990, 1.023] |
| PD | McNealis joint-RE | lag 3 | 1.001 [0.993, 1.012] | 1.016 [1.004, 1.028] | 1.001 [0.985, 1.017] | 1.000 [0.986, 1.012] |
| PD | Qu AIPW | lag 1 | 0.103 [-0.003, 0.209] | 0.118 [-0.052, 0.289] | 0.221 [0.063, 0.378] | 0.194 [0.073, 0.316] |

| | | | | | | |
|---|---|---|---|---|---|---|
| PD | Qu AIPW | lag 2 | 0.069 [-0.025, 0.163] | 0.065 [-0.055, 0.184] | 0.138 [-0.011, 0.286] | 0.113 [-0.013, 0.239] |
| PD | Qu AIPW | lag 3 | 0.066 [-0.030, 0.163] | 0.109 [-0.021, 0.239] | 0.149 [-0.020, 0.318] | 0.025 [-0.087, 0.136] |
| PD ≥4 mm | Mutual GLMM | lag 1 | 1.096 [0.816, 1.474] | 1.390 [0.998, 1.937] | 1.422 [0.854, 2.367] | 1.598 [1.036, 2.465] |
| PD ≥4 mm | Mutual GLMM | lag 2 | 1.351 [1.046, 1.744] | 1.050 [0.748, 1.473] | 1.156 [0.736, 1.817] | 0.680 [0.382, 1.211] |
| PD ≥4 mm | Mutual GLMM | lag 3 | 1.020 [0.808, 1.289] | 1.149 [0.866, 1.524] | 1.388 [0.956, 2.014] | 0.855 [0.513, 1.427] |
| PD ≥4 mm | Forastiere GPS | lag 1 | 1.627 [0.750, 3.530] | 1.304 [0.553, 3.073] | 1.970 [0.579, 6.707] | 3.866 [1.081, 13.827] |
| PD ≥4 mm | Forastiere GPS | lag 2 | 3.157 [1.409, 7.074] | 1.641 [0.591, 4.554] | 1.595 [0.384, 6.625] | 0.954 [0.171, 5.327] |
| PD ≥4 mm | Forastiere GPS | lag 3 | 2.292 [0.784, 6.695] | 3.020 [0.894, 10.200] | 2.821 [0.421, 18.892] | 1.667 [0.246, 11.311] |
| PD ≥4 mm | Qu AIPW | lag 1 | 0.015 [-0.018, 0.048] | 0.035 [-0.007, 0.076] | 0.045 [-0.002, 0.091] | 0.038 [-0.006, 0.082] |
| PD ≥4 mm | Qu AIPW | lag 2 | 0.017 [-0.012, 0.045] | 0.010 [-0.025, 0.046] | 0.037 [-0.008, 0.083] | 0.003 [-0.030, 0.036] |
| PD ≥4 mm | Qu AIPW | lag 3 | 0.009 [-0.018, 0.036] | 0.016 [-0.023, 0.056] | 0.037 [-0.008, 0.083] | 0.000 [-0.034, 0.033] |

*Abbreviations: PD = probing depth; CAL = clinical attachment level; BOP = bleeding on probing; GPS = generalized propensity score; RE = random effects; AIPW = augmented inverse-probability weighting.*

### I. Additional sensitivity analyses

Five sensitivity and falsification analyses probe the primary spillover estimate. They (i) add tooth type (anterior / premolar / molar, from the FDI position) to the outcome and propensity models; (ii) stratify the focal surfaces into approximal (mesial, distal) and smooth (buccal, palatal/lingual); (iii) sharpen the neighbourhood exposure from the neighbouring tooth as a whole to the single surface facing the focal surface; (iv) replace the baseline exposures with restorations placed between baseline and follow-up; and (v) replace the neighbourhood exposure with a negative-control exposure carried by the opposing (antagonist) tooth. All use the restoration exposure and the generalized-propensity-score machinery of Section B — individual propensity φ and neighbourhood generalized propensity score λ entered as natural cubic splines — with the same covariate set, baseline-outcome adjustment and random intercepts (person/jaw/tooth for PD; person/jaw for PD ≥4 mm), in all four waves. PD is modelled with a Gamma log-link (exp(β)) and PD ≥4 mm with a Poisson log-link (RR); estimates are posterior medians with 95% CrI.

**Supplementary Table S11.** Tooth-type sensitivity: within-mouth spillover of restorations before and after adding tooth type (anterior / premolar / molar) to the outcome and propensity models. Generalized-propensity-score mixed model (own and neighbourhood exposure) and the mutual GLMM with tooth-lags 1–3 fitted jointly; exp(β) for PD, RRs for PD ≥4 mm, with 95% CrI. Model, covariates, baseline-outcome adjustment, propensity-score terms, tooth-lags where applicable and random-intercept structure follow the main analysis (Section B).

| Wave | Outcome | Estimator | Effect | Base model | + tooth type |
|---|---|---|---|---|---|
| START-1 | PD | GPS mixed model | Direct (own) | 1.023 [1.018, 1.029] | 1.020 [1.015, 1.026] |
| START-1 | PD | GPS mixed model | Spillover (neighbouring tooth) | 1.026 [1.019, 1.033] | 1.026 [1.019, 1.034] |
| START-1 | PD | Mutual GLMM | Direct (own) | 1.018 [1.011, 1.024] | 1.012 [1.006, 1.018] |
| START-1 | PD | Mutual GLMM | Spillover, tooth-lag 1 | 1.052 [1.044, 1.059] | 1.027 [1.019, 1.034] |
| START-1 | PD | Mutual GLMM | Spillover, tooth-lag 2 | 1.017 [1.010, 1.025] | 1.005 [0.998, 1.012] |
| START-1 | PD | Mutual GLMM | Spillover, tooth-lag 3 | 0.994 [0.988, 1.000] | 1.004 [0.998, 1.011] |
| START-1 | PD ≥4 mm | GPS mixed model | Direct (own) | 1.102 [1.048, 1.158] | 1.087 [1.034, 1.143] |
| START-1 | PD ≥4 mm | GPS mixed model | Spillover (neighbouring tooth) | 1.107 [1.045, 1.172] | 1.112 [1.051, 1.178] |
| START-1 | PD ≥4 mm | Mutual GLMM | Direct (own) | 1.086 [1.024, 1.151] | 1.071 [1.010, 1.136] |
| START-1 | PD ≥4 mm | Mutual GLMM | Spillover, tooth-lag 1 | 1.217 [1.139, 1.301] | 1.096 [1.024, 1.173] |
| START-1 | PD ≥4 mm | Mutual GLMM | Spillover, tooth-lag 2 | 1.068 [1.002, 1.139] | 1.004 [0.942, 1.071] |
| START-1 | PD ≥4 mm | Mutual GLMM | Spillover, tooth-lag 3 | 0.950 [0.898, 1.006] | 0.999 [0.941, 1.061] |
| START-2 | PD | GPS mixed model | Direct (own) | 1.027 [1.021, 1.033] | 1.024 [1.018, 1.030] |
| START-2 | PD | GPS mixed model | Spillover (neighbouring tooth) | 1.032 [1.025, 1.040] | 1.032 [1.025, 1.040] |
| START-2 | PD | Mutual GLMM | Direct (own) | 1.026 [1.019, 1.033] | 1.018 [1.011, 1.024] |
| START-2 | PD | Mutual GLMM | Spillover, tooth-lag 1 | 1.079 [1.071, 1.088] | 1.036 [1.028, 1.045] |
| START-2 | PD | Mutual GLMM | Spillover, tooth-lag 2 | 1.031 [1.022, 1.039] | 1.011 [1.003, 1.018] |

| | | | | | |
|---|---|---|---|---|---|
| START-2 | PD | Mutual GLMM | Spillover, tooth-lag 3 | 0.980 [0.974, 0.987] | 1.010 [1.003, 1.017] |
| START-2 | PD ≥4 mm | GPS mixed model | Direct (own) | 1.166 [1.099, 1.238] | 1.150 [1.083, 1.221] |
| START-2 | PD ≥4 mm | GPS mixed model | Spillover (neighbouring tooth) | 1.135 [1.059, 1.217] | 1.130 [1.055, 1.211] |
| START-2 | PD ≥4 mm | Mutual GLMM | Direct (own) | 1.173 [1.095, 1.257] | 1.116 [1.043, 1.195] |
| START-2 | PD ≥4 mm | Mutual GLMM | Spillover, tooth-lag 1 | 1.330 [1.231, 1.438] | 1.098 [1.015, 1.188] |
| START-2 | PD ≥4 mm | Mutual GLMM | Spillover, tooth-lag 2 | 1.183 [1.097, 1.276] | 1.069 [0.992, 1.153] |
| START-2 | PD ≥4 mm | Mutual GLMM | Spillover, tooth-lag 3 | 0.840 [0.785, 0.899] | 0.977 [0.910, 1.050] |
| START-3 | PD | GPS mixed model | Direct (own) | 1.027 [1.019, 1.035] | 1.024 [1.016, 1.033] |
| START-3 | PD | GPS mixed model | Spillover (neighbouring tooth) | 1.044 [1.034, 1.054] | 1.041 [1.031, 1.051] |
| START-3 | PD | Mutual GLMM | Direct (own) | 1.029 [1.020, 1.038] | 1.021 [1.012, 1.030] |
| START-3 | PD | Mutual GLMM | Spillover, tooth-lag 1 | 1.081 [1.069, 1.092] | 1.041 [1.030, 1.052] |
| START-3 | PD | Mutual GLMM | Spillover, tooth-lag 2 | 1.033 [1.022, 1.044] | 1.014 [1.004, 1.025] |
| START-3 | PD | Mutual GLMM | Spillover, tooth-lag 3 | 0.987 [0.978, 0.996] | 1.012 [1.002, 1.021] |
| START-3 | PD ≥4 mm | GPS mixed model | Direct (own) | 1.224 [1.129, 1.326] | 1.228 [1.133, 1.332] |
| START-3 | PD ≥4 mm | GPS mixed model | Spillover (neighbouring tooth) | 1.245 [1.133, 1.368] | 1.225 [1.115, 1.347] |
| START-3 | PD ≥4 mm | Mutual GLMM | Direct (own) | 1.213 [1.105, 1.331] | 1.166 [1.064, 1.278] |
| START-3 | PD ≥4 mm | Mutual GLMM | Spillover, tooth-lag 1 | 1.573 [1.413, 1.752] | 1.231 [1.103, 1.372] |
| START-3 | PD ≥4 mm | Mutual GLMM | Spillover, tooth-lag 2 | 1.250 [1.129, 1.386] | 1.127 [1.018, 1.248] |
| START-3 | PD ≥4 mm | Mutual GLMM | Spillover, tooth-lag 3 | 0.886 [0.808, 0.971] | 1.058 [0.961, 1.165] |
| TREND-1 | PD | GPS mixed model | Direct (own) | 1.017 [1.012, 1.023] | 1.017 [1.011, 1.022] |
| TREND-1 | PD | GPS mixed model | Spillover (neighbouring tooth) | 1.053 [1.046, 1.060] | 1.050 [1.043, 1.057] |
| TREND-1 | PD | Mutual GLMM | Direct (own) | 1.015 [1.009, 1.021] | 1.008 [1.002, 1.014] |
| TREND-1 | PD | Mutual GLMM | Spillover, tooth-lag 1 | 1.091 [1.083, 1.098] | 1.053 [1.045, 1.060] |
| TREND-1 | PD | Mutual GLMM | Spillover, tooth-lag 2 | 1.029 [1.022, 1.036] | 1.015 [1.008, 1.022] |
| TREND-1 | PD | Mutual GLMM | Spillover, tooth-lag 3 | 0.993 [0.987, 0.999] | 1.015 [1.008, 1.021] |
| TREND-1 | PD ≥4 mm | GPS mixed model | Direct (own) | 1.109 [1.043, 1.179] | 1.106 [1.040, 1.175] |
| TREND-1 | PD ≥4 mm | GPS mixed model | Spillover (neighbouring tooth) | 1.227 [1.141, 1.318] | 1.219 [1.134, 1.310] |
| TREND-1 | PD ≥4 mm | Mutual GLMM | Direct (own) | 1.106 [1.033, 1.183] | 1.046 [0.977, 1.119] |
| TREND-1 | PD ≥4 mm | Mutual GLMM | Spillover, tooth-lag 1 | 1.506 [1.391, 1.630] | 1.202 [1.108, 1.305] |
| TREND-1 | PD ≥4 mm | Mutual GLMM | Spillover, tooth-lag 2 | 1.132 [1.049, 1.221] | 1.052 [0.976, 1.135] |
| TREND-1 | PD ≥4 mm | Mutual GLMM | Spillover, tooth-lag 3 | 0.968 [0.904, 1.036] | 1.086 [1.011, 1.167] |

*Analytic samples are identical within an estimator; only the linear predictor and propensity models differ. Abbreviations: PD = probing depth; CrI = credible interval; GPS = generalized propensity score; RR = risk ratio.*

**Supplementary Table S12.** Surface-stratum sensitivity: the generalized-propensity-score spillover fitted separately in approximal (mesial, distal) and smooth (buccal, palatal/lingual) focal surfaces. The neighbourhood exposure remains tooth-level and is built on all surfaces before stratification; the propensity and generalized propensity scores are re-estimated within each stratum. exp(β) for PD, RRs for PD ≥4 mm, with 95% CrI. Model, covariates, baseline-outcome adjustment, propensity-score terms, tooth-lags where applicable and random-intercept structure follow the main analysis (Section B).

| Wave | Outcome | Effect | Approximal focal surfaces | Smooth focal surfaces |
|---|---|---|---|---|
| START-1 | PD | Direct (own) | 1.011 [1.004, 1.017] | 1.045 [1.036, 1.055] |
| START-1 | PD | Spillover (neighbouring tooth) | 1.025 [1.016, 1.033] | 1.037 [1.027, 1.047] |
| START-1 | PD ≥4 mm | Direct (own) | 1.103 [1.044, 1.165] | 1.103 [0.980, 1.241] |
| START-1 | PD ≥4 mm | Spillover (neighbouring tooth) | 1.104 [1.036, 1.176] | 1.129 [0.995, 1.281] |
| START-2 | PD | Direct (own) | 1.014 [1.007, 1.021] | 1.068 [1.058, 1.079] |
| START-2 | PD | Spillover (neighbouring tooth) | 1.031 [1.023, 1.040] | 1.055 [1.044, 1.067] |
| START-2 | PD ≥4 mm | Direct (own) | 1.122 [1.053, 1.195] | 1.239 [1.064, 1.443] |
| START-2 | PD ≥4 mm | Spillover (neighbouring tooth) | 1.140 [1.059, 1.228] | 1.187 [1.007, 1.399] |
| START-3 | PD | Direct (own) | 1.017 [1.008, 1.027] | 1.044 [1.031, 1.058] |
| START-3 | PD | Spillover (neighbouring tooth) | 1.041 [1.029, 1.053] | 1.058 [1.043, 1.073] |
| START-3 | PD ≥4 mm | Direct (own) | 1.220 [1.119, 1.329] | 1.092 [0.889, 1.341] |
| START-3 | PD ≥4 mm | Spillover (neighbouring tooth) | 1.237 [1.118, 1.369] | 1.305 [1.045, 1.631] |
| TREND-1 | PD | Direct (own) | 1.008 [1.001, 1.015] | 1.054 [1.045, 1.062] |
| TREND-1 | PD | Spillover (neighbouring tooth) | 1.031 [1.022, 1.039] | 1.084 [1.074, 1.094] |
| TREND-1 | PD ≥4 mm | Direct (own) | 1.093 [1.022, 1.169] | 1.294 [1.135, 1.476] |
| TREND-1 | PD ≥4 mm | Spillover (neighbouring tooth) | 1.190 [1.099, 1.287] | 1.317 [1.138, 1.524] |

*Analysed surfaces (approximal/smooth): START-1 49738/37859; START-2 37167/28251; START-3 26509/20158; TREND-1 44089/44047. SHIP-START-0 did not probe the mandibular lingual sites, so the SHIP-START smooth stratum is almost entirely buccal plus maxillary palatal and is not a balanced buccal/lingual contrast; SHIP-TREND-1 has complete coverage and is the wave to read for this comparison. Abbreviations: PD = probing depth; CrI = credible interval; RR = risk ratio.*

**Supplementary Table S13.** Contact-specific neighbourhood exposure: the neighbourhood exposure sharpened from the neighbouring tooth as a whole to the single surface of the neighbouring tooth facing the focal surface (focal mesial ↔ neighbour distal; focal distal ↔ neighbour mesial). Three generalized-propensity-score models per wave and outcome — facing surface only, neighbouring tooth only (the like-for-like comparator in the same sample), and both together. exp(β) for PD, RRs for PD ≥4 mm, with 95% CrI. Model, covariates, baseline-outcome adjustment, propensity-score terms, tooth-lags where applicable and random-intercept structure follow the main analysis (Section B).

| Wave | Outcome | Neighbourhood exposure | Effect | Estimate [95% CrI] |
|---|---|---|---|---|
| START-1 | PD | Facing surface only | Direct (own) | 0.975 [0.961, 0.990] |
| START-1 | PD | Facing surface only | Spillover (facing surface) | 0.993 [0.983, 1.003] |
| START-1 | PD | Neighbouring tooth only | Direct (own) | 1.020 [1.013, 1.028] |
| START-1 | PD | Neighbouring tooth only | Spillover (neighbouring tooth) | 1.008 [0.999, 1.018] |
| START-1 | PD | Both exposures | Direct (own) | 0.981 [0.966, 0.997] |
| START-1 | PD | Both exposures | Spillover (neighbouring tooth) | 1.002 [0.991, 1.013] |
| START-1 | PD | Both exposures | Spillover (facing surface) | 0.994 [0.983, 1.005] |
| START-1 | PD ≥4 mm | Facing surface only | Direct (own) | 0.885 [0.781, 1.004] |
| START-1 | PD ≥4 mm | Facing surface only | Spillover (facing surface) | 0.937 [0.858, 1.023] |
| START-1 | PD ≥4 mm | Neighbouring tooth only | Direct (own) | 1.181 [1.107, 1.260] |
| START-1 | PD ≥4 mm | Neighbouring tooth only | Spillover (neighbouring tooth) | 1.041 [0.965, 1.124] |
| START-1 | PD ≥4 mm | Both exposures | Direct (own) | 0.919 [0.803, 1.052] |
| START-1 | PD ≥4 mm | Both exposures | Spillover (neighbouring tooth) | 1.015 [0.925, 1.114] |
| START-1 | PD ≥4 mm | Both exposures | Spillover (facing surface) | 0.944 [0.855, 1.043] |
| START-2 | PD | Facing surface only | Direct (own) | 1.009 [0.992, 1.027] |
| START-2 | PD | Facing surface only | Spillover (facing surface) | 1.013 [1.002, 1.024] |
| START-2 | PD | Neighbouring tooth only | Direct (own) | 1.019 [1.011, 1.026] |
| START-2 | PD | Neighbouring tooth only | Spillover (neighbouring tooth) | 1.020 [1.011, 1.030] |
| START-2 | PD | Both exposures | Direct (own) | 1.019 [1.001, 1.038] |
| START-2 | PD | Both exposures | Spillover (neighbouring tooth) | 1.006 [0.995, 1.017] |
| START-2 | PD | Both exposures | Spillover (facing surface) | 1.015 [1.003, 1.028] |
| START-2 | PD ≥4 mm | Facing surface only | Direct (own) | 0.974 [0.838, 1.132] |
| START-2 | PD ≥4 mm | Facing surface only | Spillover (facing surface) | 1.011 [0.912, 1.121] |
| START-2 | PD ≥4 mm | Neighbouring tooth only | Direct (own) | 1.170 [1.086, 1.261] |
| START-2 | PD ≥4 mm | Neighbouring tooth only | Spillover (neighbouring tooth) | 1.102 [1.009, 1.203] |
| START-2 | PD ≥4 mm | Both exposures | Direct (own) | 1.050 [0.892, 1.237] |
| START-2 | PD ≥4 mm | Both exposures | Spillover (neighbouring tooth) | 1.040 [0.934, 1.159] |
| START-2 | PD ≥4 mm | Both exposures | Spillover (facing surface) | 1.028 [0.914, 1.157] |
| START-3 | PD | Facing surface only | Direct (own) | 0.994 [0.971, 1.017] |
| START-3 | PD | Facing surface only | Spillover (facing surface) | 1.003 [0.987, 1.019] |
| START-3 | PD | Neighbouring tooth only | Direct (own) | 1.024 [1.013, 1.035] |
| START-3 | PD | Neighbouring tooth only | Spillover (neighbouring tooth) | 1.027 [1.014, 1.040] |
| START-3 | PD | Both exposures | Direct (own) | 1.007 [0.982, 1.032] |
| START-3 | PD | Both exposures | Spillover (neighbouring tooth) | 1.013 [0.998, 1.028] |
| START-3 | PD | Both exposures | Spillover (facing surface) | 1.005 [0.987, 1.023] |

| | | | | |
|---|---|---|---|---|
| START-3 | PD ≥4 mm | Facing surface only | Direct (own) | 0.782 [0.637, 0.960] |
| START-3 | PD ≥4 mm | Facing surface only | Spillover (facing surface) | 0.927 [0.801, 1.072] |
| START-3 | PD ≥4 mm | Neighbouring tooth only | Direct (own) | 1.284 [1.160, 1.421] |
| START-3 | PD ≥4 mm | Neighbouring tooth only | Spillover (neighbouring tooth) | 1.214 [1.076, 1.369] |
| START-3 | PD ≥4 mm | Both exposures | Direct (own) | 0.909 [0.728, 1.135] |
| START-3 | PD ≥4 mm | Both exposures | Spillover (neighbouring tooth) | 1.094 [0.944, 1.267] |
| START-3 | PD ≥4 mm | Both exposures | Spillover (facing surface) | 0.964 [0.818, 1.136] |
| TREND-1 | PD | Facing surface only | Direct (own) | 1.007 [0.996, 1.019] |
| TREND-1 | PD | Facing surface only | Spillover (facing surface) | 0.999 [0.991, 1.008] |
| TREND-1 | PD | Neighbouring tooth only | Direct (own) | 1.016 [1.009, 1.024] |
| TREND-1 | PD | Neighbouring tooth only | Spillover (neighbouring tooth) | 1.018 [1.009, 1.028] |
| TREND-1 | PD | Both exposures | Direct (own) | 1.011 [0.999, 1.023] |
| TREND-1 | PD | Both exposures | Spillover (neighbouring tooth) | 1.018 [1.007, 1.029] |
| TREND-1 | PD | Both exposures | Spillover (facing surface) | 0.995 [0.985, 1.005] |
| TREND-1 | PD ≥4 mm | Facing surface only | Direct (own) | 1.011 [0.906, 1.127] |
| TREND-1 | PD ≥4 mm | Facing surface only | Spillover (facing surface) | 0.982 [0.898, 1.074] |
| TREND-1 | PD ≥4 mm | Neighbouring tooth only | Direct (own) | 1.136 [1.049, 1.231] |
| TREND-1 | PD ≥4 mm | Neighbouring tooth only | Spillover (neighbouring tooth) | 1.151 [1.046, 1.267] |
| TREND-1 | PD ≥4 mm | Both exposures | Direct (own) | 1.026 [0.914, 1.151] |
| TREND-1 | PD ≥4 mm | Both exposures | Spillover (neighbouring tooth) | 1.127 [1.003, 1.266] |
| TREND-1 | PD ≥4 mm | Both exposures | Spillover (facing surface) | 0.942 [0.851, 1.043] |

*Restricted to approximal focal surfaces whose facing partner tooth is present (START-1 39665, START-2 29619, START-3 21165, TREND-1 35344 surfaces), so the estimates are not comparable with the full-sample analysis. The two neighbourhood exposures are strongly correlated, so the 'both exposures' model is a direction-of-effect check rather than two independent estimates. Abbreviations: PD = probing depth; CrI = credible interval; RR = risk ratio.*

**Supplementary Table S14.** Incident-restoration sensitivity: the own and neighbourhood exposures redefined as restorations placed between the baseline and the follow-up examination among baseline-sound surfaces, fitted alone and with the baseline neighbourhood exposure retained in the same model. Generalized-propensity-score mixed model; exp(β) for PD, RRs for PD ≥4 mm, with 95% CrI. Model, covariates, baseline-outcome adjustment, propensity-score terms, tooth-lags where applicable and random-intercept structure follow the main analysis (Section B).

| Wave | Outcome | Model | Effect | Estimate [95% CrI] |
|---|---|---|---|---|
| START-1 | PD | Incident exposures only | Direct (incident own restoration) | 1.025 [1.014, 1.036] |
| START-1 | PD | Incident exposures only | Spillover (incident neighbouring restoration) | 0.983 [0.961, 1.006] |
| START-1 | PD | Incident + baseline neighbourhood | Direct (incident own restoration) | 1.022 [1.012, 1.033] |
| START-1 | PD | Incident + baseline neighbourhood | Spillover (incident neighbouring restoration) | 0.983 [0.960, 1.005] |
| START-1 | PD | Incident + baseline neighbourhood | Spillover (baseline neighbourhood) | 1.031 [1.023, 1.040] |
| START-1 | PD ≥4 mm | Incident exposures only | Direct (incident own restoration) | 1.107 [1.005, 1.221] |
| START-1 | PD ≥4 mm | Incident exposures only | Spillover (incident neighbouring restoration) | 0.920 [0.750, 1.127] |
| START-1 | PD ≥4 mm | Incident + baseline neighbourhood | Direct (incident own restoration) | 1.093 [0.991, 1.205] |
| START-1 | PD ≥4 mm | Incident + baseline neighbourhood | Spillover (incident neighbouring restoration) | 0.918 [0.749, 1.125] |
| START-1 | PD ≥4 mm | Incident + baseline neighbourhood | Spillover (baseline neighbourhood) | 1.117 [1.041, 1.198] |
| START-2 | PD | Incident exposures only | Direct (incident own restoration) | 1.017 [1.005, 1.028] |
| START-2 | PD | Incident exposures only | Spillover (incident neighbouring restoration) | 0.999 [0.984, 1.015] |
| START-2 | PD | Incident + baseline neighbourhood | Direct (incident own restoration) | 1.013 [1.002, 1.024] |
| START-2 | PD | Incident + baseline neighbourhood | Spillover (incident neighbouring restoration) | 0.997 [0.982, 1.013] |
| START-2 | PD | Incident + baseline neighbourhood | Spillover (baseline neighbourhood) | 1.035 [1.026, 1.043] |
| START-2 | PD ≥4 mm | Incident exposures only | Direct (incident own restoration) | 1.063 [0.945, 1.195] |
| START-2 | PD ≥4 mm | Incident exposures only | Spillover (incident neighbouring restoration) | 0.876 [0.755, 1.016] |
| START-2 | PD ≥4 mm | Incident + baseline neighbourhood | Direct (incident own restoration) | 1.052 [0.935, 1.183] |
| START-2 | PD ≥4 mm | Incident + baseline neighbourhood | Spillover (incident neighbouring restoration) | 0.875 [0.754, 1.015] |
| START-2 | PD ≥4 mm | Incident + baseline neighbourhood | Spillover (baseline neighbourhood) | 1.112 [1.021, 1.211] |
| START-3 | PD | Incident exposures only | Direct (incident own restoration) | 1.011 [0.996, 1.027] |
| START-3 | PD | Incident exposures only | Spillover (incident neighbouring restoration) | 0.994 [0.978, 1.010] |
| START-3 | PD | Incident + baseline neighbourhood | Direct (incident own restoration) | 1.008 [0.993, 1.023] |

| | | | | |
|---|---|---|---|---|
| START-3 | PD | Incident + baseline neighbourhood | Spillover (incident neighbouring restoration) | 0.991 [0.976, 1.007] |
| START-3 | PD | Incident + baseline neighbourhood | Spillover (baseline neighbourhood) | 1.050 [1.039, 1.062] |
| START-3 | PD ≥4 mm | Incident exposures only | Direct (incident own restoration) | 0.902 [0.766, 1.061] |
| START-3 | PD ≥4 mm | Incident exposures only | Spillover (incident neighbouring restoration) | 0.909 [0.779, 1.061] |
| START-3 | PD ≥4 mm | Incident + baseline neighbourhood | Direct (incident own restoration) | 0.894 [0.760, 1.053] |
| START-3 | PD ≥4 mm | Incident + baseline neighbourhood | Spillover (incident neighbouring restoration) | 0.899 [0.770, 1.050] |
| START-3 | PD ≥4 mm | Incident + baseline neighbourhood | Spillover (baseline neighbourhood) | 1.332 [1.183, 1.499] |
| TREND-1 | PD | Incident exposures only | Direct (incident own restoration) | 1.019 [1.009, 1.030] |
| TREND-1 | PD | Incident exposures only | Spillover (incident neighbouring restoration) | 1.004 [0.978, 1.030] |
| TREND-1 | PD | Incident + baseline neighbourhood | Direct (incident own restoration) | 1.016 [1.005, 1.026] |
| TREND-1 | PD | Incident + baseline neighbourhood | Spillover (incident neighbouring restoration) | 1.002 [0.976, 1.028] |
| TREND-1 | PD | Incident + baseline neighbourhood | Spillover (baseline neighbourhood) | 1.042 [1.034, 1.050] |
| TREND-1 | PD ≥4 mm | Incident exposures only | Direct (incident own restoration) | 0.991 [0.875, 1.123] |
| TREND-1 | PD ≥4 mm | Incident exposures only | Spillover (incident neighbouring restoration) | 0.906 [0.676, 1.213] |
| TREND-1 | PD ≥4 mm | Incident + baseline neighbourhood | Direct (incident own restoration) | 0.972 [0.858, 1.101] |
| TREND-1 | PD ≥4 mm | Incident + baseline neighbourhood | Spillover (incident neighbouring restoration) | 0.898 [0.671, 1.204] |
| TREND-1 | PD ≥4 mm | Incident + baseline neighbourhood | Spillover (baseline neighbourhood) | 1.221 [1.115, 1.338] |

*Incident exposures are ascertained at the follow-up examination and are therefore contemporaneous with the outcome; reverse causation cannot be excluded. The incident own-restoration effect is positive at the shorter follow-ups and null by SHIP-START-3, whereas the incident neighbourhood effect is null throughout and the baseline neighbourhood effect persists in the same model. Abbreviations: PD = probing depth; CrI = credible interval; RR = risk ratio.*

**Supplementary Table S15.** Negative-control exposure carried by the opposing (antagonist) tooth — same side, opposite jaw, same position — fitted in place of, and alongside, the neighbourhood exposure. SHIP examines a randomly selected half-mouth (quadrants 1+4 or 2+3), so the contralateral mirror tooth lies in the unexamined half and its restoration status is recorded for 0.0% of teeth; the opposing tooth is the feasible mirror, available for START-1 82.4%, START-2 83.0%, START-3 83.2%, TREND-1 86.2% of teeth. It shares person-, side-, examiner- and care-seeking-level confounding with the focal tooth but has no interproximal contact with it, so a non-null coefficient would indicate residual mouth-level confounding rather than local interference. exp(β) for PD, RRs for PD ≥4 mm, with 95% CrI. Model, covariates, baseline-outcome adjustment, propensity-score terms, tooth-lags where applicable and random-intercept structure follow the main analysis (Section B).

| Wave | Outcome | Model | Effect | Estimate [95% CrI] |
|---|---|---|---|---|
| START-1 | PD | Negative control only | Direct (own) | 1.025 [1.019, 1.031] |
| START-1 | PD | Negative control only | Negative control (opposing tooth) | 0.981 [0.975, 0.988] |
| START-1 | PD | Negative control + neighbourhood | Direct (own) | 1.026 [1.020, 1.032] |
| START-1 | PD | Negative control + neighbourhood | Spillover (neighbouring tooth) | 1.026 [1.018, 1.034] |
| START-1 | PD | Negative control + neighbourhood | Negative control (opposing tooth) | 0.986 [0.979, 0.993] |
| START-1 | PD ≥4 mm | Negative control only | Direct (own) | 1.133 [1.073, 1.196] |
| START-1 | PD ≥4 mm | Negative control only | Negative control (opposing tooth) | 0.958 [0.905, 1.014] |
| START-1 | PD ≥4 mm | Negative control + neighbourhood | Direct (own) | 1.128 [1.065, 1.195] |
| START-1 | PD ≥4 mm | Negative control + neighbourhood | Spillover (neighbouring tooth) | 1.128 [1.055, 1.206] |
| START-1 | PD ≥4 mm | Negative control + neighbourhood | Negative control (opposing tooth) | 0.985 [0.929, 1.044] |
| START-2 | PD | Negative control only | Direct (own) | 1.030 [1.024, 1.037] |
| START-2 | PD | Negative control only | Negative control (opposing tooth) | 0.978 [0.971, 0.985] |
| START-2 | PD | Negative control + neighbourhood | Direct (own) | 1.031 [1.024, 1.038] |
| START-2 | PD | Negative control + neighbourhood | Spillover (neighbouring tooth) | 1.035 [1.027, 1.043] |
| START-2 | PD | Negative control + neighbourhood | Negative control (opposing tooth) | 0.984 [0.977, 0.991] |
| START-2 | PD ≥4 mm | Negative control only | Direct (own) | 1.179 [1.106, 1.256] |
| START-2 | PD ≥4 mm | Negative control only | Negative control (opposing tooth) | 0.871 [0.814, 0.931] |
| START-2 | PD ≥4 mm | Negative control + neighbourhood | Direct (own) | 1.197 [1.119, 1.281] |
| START-2 | PD ≥4 mm | Negative control + neighbourhood | Spillover (neighbouring tooth) | 1.130 [1.044, 1.223] |
| START-2 | PD ≥4 mm | Negative control + neighbourhood | Negative control (opposing tooth) | 0.905 [0.845, 0.969] |
| START-3 | PD | Negative control only | Direct (own) | 1.032 [1.023, 1.041] |
| START-3 | PD | Negative control only | Negative control (opposing tooth) | 0.976 [0.966, 0.985] |
| START-3 | PD | Negative control + neighbourhood | Direct (own) | 1.030 [1.021, 1.039] |
| START-3 | PD | Negative control + neighbourhood | Spillover (neighbouring tooth) | 1.043 [1.032, 1.054] |
| START-3 | PD | Negative control + neighbourhood | Negative control (opposing tooth) | 0.982 [0.972, 0.991] |
| START-3 | PD ≥4 mm | Negative control only | Direct (own) | 1.252 [1.146, 1.368] |
| START-3 | PD ≥4 mm | Negative control only | Negative control (opposing tooth) | 0.867 [0.789, 0.953] |
| START-3 | PD ≥4 mm | Negative control + neighbourhood | Direct (own) | 1.276 [1.164, 1.400] |
| START-3 | PD ≥4 mm | Negative control + neighbourhood | Spillover (neighbouring tooth) | 1.222 [1.096, 1.361] |

| | | | | |
|---|---|---|---|---|
| START-3 | PD ≥4 mm | Negative control + neighbourhood | Negative control (opposing tooth) | 0.915 [0.831, 1.007] |
| TREND-1 | PD | Negative control only | Direct (own) | 1.024 [1.018, 1.029] |
| TREND-1 | PD | Negative control only | Negative control (opposing tooth) | 0.974 [0.968, 0.980] |
| TREND-1 | PD | Negative control + neighbourhood | Direct (own) | 1.020 [1.014, 1.026] |
| TREND-1 | PD | Negative control + neighbourhood | Spillover (neighbouring tooth) | 1.054 [1.046, 1.061] |
| TREND-1 | PD | Negative control + neighbourhood | Negative control (opposing tooth) | 0.982 [0.976, 0.988] |
| TREND-1 | PD ≥4 mm | Negative control only | Direct (own) | 1.138 [1.066, 1.215] |
| TREND-1 | PD ≥4 mm | Negative control only | Negative control (opposing tooth) | 0.845 [0.792, 0.902] |
| TREND-1 | PD ≥4 mm | Negative control + neighbourhood | Direct (own) | 1.139 [1.063, 1.221] |
| TREND-1 | PD ≥4 mm | Negative control + neighbourhood | Spillover (neighbouring tooth) | 1.245 [1.147, 1.352] |
| TREND-1 | PD ≥4 mm | Negative control + neighbourhood | Negative control (opposing tooth) | 0.889 [0.832, 0.951] |

*Restricted to focal teeth with an opposing partner present, so the sample is smaller than the main analysis. Opposing teeth are in occlusal contact, so a restoration there could in principle alter loading; the control is therefore conservative for occlusion but clean for interproximal ecology. Abbreviations: PD = probing depth; CrI = credible interval; RR = risk ratio.*

### J. Confirmatory analysis: untreated caries

*The restoration analysis was repeated with untreated caries as the focal exposure, under the identical covariate set and pipeline. Restored and missing surfaces were excluded so that the two exposures are disjoint. Untreated caries is rare in this well-restored population (0.1–0.3% of surfaces at baseline; Table S1), so estimates are less precise and weighting leaves more residual imbalance than for restorations.*

**Supplementary Table S16.** Direct effect of untreated caries on the periodontal site at the affected surface (Model A: baseline status; Model B: incident caries). GLMM (Gamma-log PD/CAL, Poisson-log BOP/PD ≥4 mm; person/jaw/tooth random intercepts); exp(β) for PD/CAL, RRs for BOP/PD ≥4 mm, 95% CrI.

| Model | Outcome | Category | START-1 | START-2 | START-3 | TREND-1 |
|---|---|---|---|---|---|---|
| A: baseline status | BOP | Caries | 1.110 [0.887, 1.390] | 1.221 [0.944, 1.580] | 1.022 [0.722, 1.447] | 0.772 [0.484, 1.233] |
| A: baseline status | CAL | Caries | 1.092 [0.999, 1.193] | 1.066 [0.979, 1.161] | 1.102 [0.991, 1.227] | 1.071 [0.971, 1.181] |
| A: baseline status | PD | Caries | 1.024 [0.997, 1.051] | 1.034 [1.003, 1.065] | 1.072 [1.028, 1.119] | 1.015 [0.972, 1.059] |
| A: baseline status | PD ≥4 mm | Caries | 1.032 [0.838, 1.271] | 1.227 [0.957, 1.574] | 1.571 [1.099, 2.244] | 1.153 [0.795, 1.672] |
| B: incident | BOP | Caries (incident) | 0.958 [0.608, 1.511] | 1.191 [0.779, 1.821] | 1.323 [0.888, 1.970] | 1.395 [0.769, 2.530] |
| B: incident | CAL | Caries (incident) | 1.223 [1.066, 1.404] | 1.136 [0.999, 1.291] | 0.975 [0.857, 1.109] | 1.009 [0.886, 1.148] |
| B: incident | PD | Caries (incident) | 1.025 [0.980, 1.072] | 1.058 [1.007, 1.112] | 0.987 [0.934, 1.042] | 0.982 [0.925, 1.043] |
| B: incident | PD ≥4 mm | Caries (incident) | 1.036 [0.765, 1.404] | 1.338 [0.965, 1.855] | 0.810 [0.500, 1.314] | 0.783 [0.458, 1.338] |

*Abbreviations: PD = probing depth; CAL = clinical attachment level; BOP = bleeding on probing; CrI = credible interval; RR = risk ratio.*

**Supplementary Table S17.** Spillover from untreated caries on neighbouring teeth: neighbourhood-exposure mixed model with individual and neighbourhood generalized-propensity-score adjustment; direct (own) and spillover (neighbour) effects.

| Outcome | Scale | Effect | START-1 | START-2 | START-3 | TREND-1 |
|---|---|---|---|---|---|---|
| BOP | RR | Direct (own) | 1.216 [0.802, 1.843] | 1.072 [0.624, 1.839] | 1.165 [0.601, 2.260] | 0.234 [0.076, 0.718] |
| BOP | RR | Spillover (neighbour) | 1.027 [0.576, 1.833] | 1.346 [0.631, 2.873] | 1.932 [0.822, 4.541] | 1.958 [0.604, 6.346] |
| CAL | ratio | Direct (own) | 1.355 [1.137, 1.616] | 0.886 [0.759, 1.035] | 1.073 [0.899, 1.280] | 0.835 [0.706, 0.987] |
| CAL | ratio | Spillover (neighbour) | 0.958 [0.759, 1.209] | 1.322 [1.084, 1.613] | 1.007 [0.800, 1.266] | 1.516 [1.221, 1.882] |
| PD | ratio | Direct (own) | 1.084 [1.034, 1.137] | 0.876 [0.829, 0.925] | 0.994 [0.924, 1.070] | 0.864 [0.798, 0.937] |
| PD | ratio | Spillover (neighbour) | 0.974 [0.914, 1.038] | 1.106 [1.025, 1.194] | 1.012 [0.912, 1.122] | 1.171 [1.052, 1.303] |
| PD ≥4 mm | RR | Direct (own) | 1.319 [0.908, 1.914] | 0.574 [0.340, 0.970] | 0.653 [0.289, 1.473] | 0.339 [0.147, 0.777] |
| PD ≥4 mm | RR | Spillover (neighbour) | 0.961 [0.570, 1.620] | 1.264 [0.641, 2.491] | 1.214 [0.431, 3.422] | 3.544 [1.349, 9.309] |

*Abbreviations: PD = probing depth; CAL = clinical attachment level; BOP = bleeding on probing; RR = risk ratio.*

**Supplementary Table S18.** Neighbouring-tooth caries effect on PD and CAL across three estimators (generalized-propensity-score mixed model; joint correlated-random-effects model, indirect effect; doubly-robust AIPW, average spillover effect). 95% CrI for the two Bayesian models; 95% CI for AIPW.

| Outcome | Estimator | START-1 | START-2 | START-3 | TREND-1 |
|---|---|---|---|---|---|
| CAL | Doubly-robust AIPW (ASE, mm) | 0.133 [-0.064, 0.331] | 0.209 [-0.011, 0.429] | 0.043 [-0.263, 0.349] | 0.045 [-0.174, 0.264] |
| CAL | GPS GLMM (neighbour, ratio) | 0.958 [0.759, 1.209] | 1.322 [1.084, 1.613] | 1.007 [0.800, 1.266] | 1.516 [1.221, 1.882] |
| CAL | Joint correlated-RE (IE, ratio) | 0.964 [0.927, 1.007] | 0.994 [0.951, 1.041] | 0.971 [0.919, 1.026] | 1.027 [0.990, 1.069] |
| PD | Doubly-robust AIPW (ASE, mm) | 0.084 [-0.007, 0.175] | 0.159 [0.034, 0.284] | 0.133 [-0.010, 0.276] | 0.234 [0.108, 0.360] |
| PD | GPS GLMM (neighbour, ratio) | 0.974 [0.914, 1.038] | 1.106 [1.025, 1.194] | 1.012 [0.912, 1.122] | 1.171 [1.052, 1.303] |
| PD | Joint correlated-RE (IE, ratio) | 1.007 [0.996, 1.018] | 1.012 [0.997, 1.026] | 1.004 [0.985, 1.022] | 1.035 [1.018, 1.051] |

*Abbreviations: PD = probing depth; CAL = clinical attachment level; CrI = credible interval; CI = confidence interval; GPS = generalized propensity score; RE = random effects; AIPW = augmented inverse-probability weighting.*

**Supplementary Table S19.** Baseline covariates by own caries status (untreated caries vs. sound) before and after inverse-probability-of-treatment weighting, SHIP-START. Values are the mean (SD) of each continuous covariate and the percentage in each category of the categorical covariates, given separately for carious and for sound surfaces, together with the standardized mean difference (SMD) between the two groups before and after weighting.

| Variable | Sound | Caries | SMD | Sound (IPTW) | Caries (IPTW) | SMD (IPTW) |
|---|---|---|---|---|---|---|
| Age, yr | 42.6 (13.6) | 43.5 (14.1) | 0.062 | 42.6 (13.6) | 42.9 (13.7) | 0.018 |
| Male, % | 50.7 | 51.0 | 0.005 | 50.7 | 50.9 | 0.004 |
| School <10 yr, % | 21.4 | 29.4 | 0.184 | 21.5 | 25.3 | 0.089 |
| School 10 yr, % | 56.5 | 50.6 | -0.118 | 56.4 | 53.2 | -0.064 |
| School >10 yr, % | 22.1 | 20.0 | -0.051 | 22.0 | 21.5 | -0.013 |
| Never smoker, % | 37.3 | 30.3 | -0.149 | 37.3 | 34.0 | -0.068 |
| Former smoker, % | 32.9 | 32.7 | -0.003 | 32.9 | 32.3 | -0.012 |
| Current smoker, % | 29.8 | 37.0 | 0.152 | 29.8 | 33.6 | 0.082 |
| Pack-years | 8.4 (11.2) | 9.4 (11.7) | 0.087 | 8.4 (11.2) | 8.8 (11.7) | 0.038 |
| BMI, kg/m2 | 26.6 (4.6) | 26.7 (4.6) | 0.024 | 26.6 (4.6) | 26.5 (4.6) | -0.011 |
| Diabetes, % | 5.3 | 7.0 | 0.070 | 5.3 | 6.0 | 0.031 |
| Toothbrushing >=2x/day, % | 84.3 | 79.3 | -0.130 | 84.3 | 82.7 | -0.043 |
| Interdental care, % | 39.4 | 28.5 | -0.230 | 39.3 | 33.9 | -0.112 |
| Dental visit <=12 mo, % | 89.6 | 80.4 | -0.261 | 89.6 | 87.0 | -0.079 |

*Surface-level values; subject-level characteristics in Table S1. Abbreviations: IPTW = inverse-probability-of-treatment weighting; SMD = standardized mean difference.*

**Supplementary Table S20.** Baseline covariates by own caries status before and after inverse-probability-of-treatment weighting, SHIP-TREND. Values are the mean (SD) of each continuous covariate and the percentage in each category of the categorical covariates, given separately for carious and for sound surfaces, together with the standardized mean difference (SMD) between the two groups before and after weighting.

| Variable | Sound | Caries | SMD | Sound (IPTW) | Caries (IPTW) | SMD (IPTW) |
|---|---|---|---|---|---|---|
| Age, yr | 44.9 (13.2) | 45.8 (14.4) | 0.066 | 44.9 (13.2) | 44.8 (14.0) | -0.012 |
| Male, % | 50.8 | 56.2 | 0.109 | 50.8 | 54.7 | 0.078 |
| School <10 yr, % | 8.8 | 17.8 | 0.267 | 8.9 | 10.6 | 0.058 |
| School 10 yr, % | 55.5 | 53.0 | -0.051 | 55.5 | 56.6 | 0.022 |
| School >10 yr, % | 35.6 | 29.2 | -0.138 | 35.6 | 32.8 | -0.059 |
| Never smoker, % | 39.1 | 36.2 | -0.059 | 39.1 | 39.8 | 0.015 |
| Former smoker, % | 36.8 | 23.8 | -0.287 | 36.8 | 31.2 | -0.119 |
| Current smoker, % | 24.1 | 40.0 | 0.346 | 24.1 | 29.0 | 0.111 |
| Pack-years | 8.1 (10.4) | 10.3 (11.8) | 0.199 | 8.1 (10.5) | 8.4 (10.2) | 0.029 |
| BMI, kg/m2 | 27.0 (4.5) | 27.5 (5.0) | 0.103 | 27.0 (4.5) | 27.6 (4.9) | 0.131 |
| Diabetes, % | 5.7 | 5.4 | -0.011 | 5.7 | 5.4 | -0.010 |
| Toothbrushing >=2x/day, % | 86.9 | 79.5 | -0.199 | 86.9 | 84.4 | -0.071 |
| Interdental care, % | 22.8 | 20.5 | -0.056 | 22.8 | 22.0 | -0.019 |
| Dental visit <=12 mo, % | 91.2 | 77.8 | -0.375 | 91.1 | 86.4 | -0.150 |

*Surface-level values; subject-level characteristics in Table S1. Abbreviations: IPTW = inverse-probability-of-treatment weighting; SMD = standardized mean difference.*

**Supplementary Figure S3.** Direct and spillover effects of untreated caries on periodontal status (neighbourhood-exposure generalized-propensity-score models), across SHIP-START follow-ups and the SHIP-TREND replication. Confirmatory exposure.

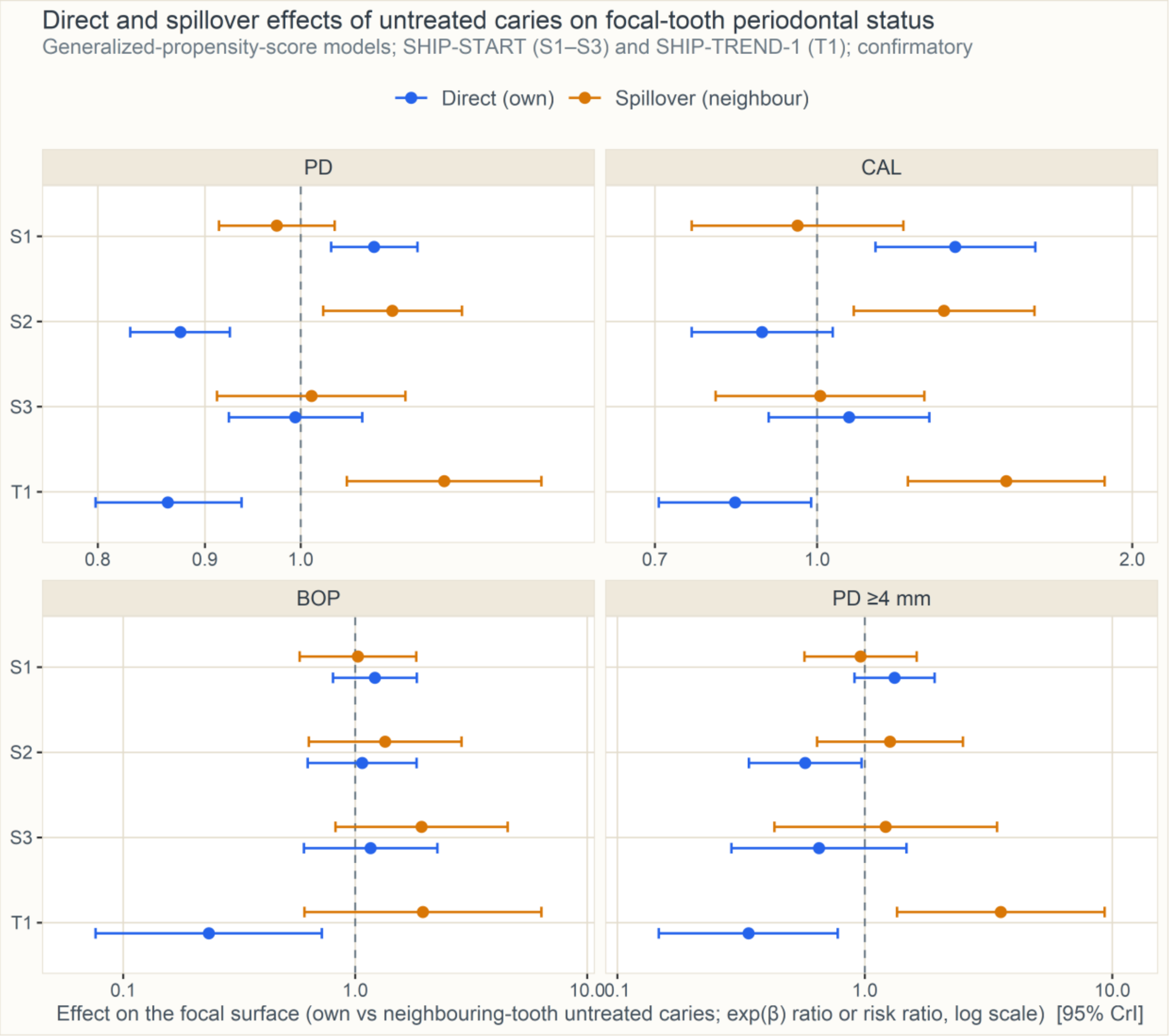

**Supplementary Figure S4.** Neighbouring-tooth caries effect on PD across three estimators (generalized-propensity-score mixed model, joint correlated-random-effects model, doubly-robust AIPW). Confirmatory exposure.

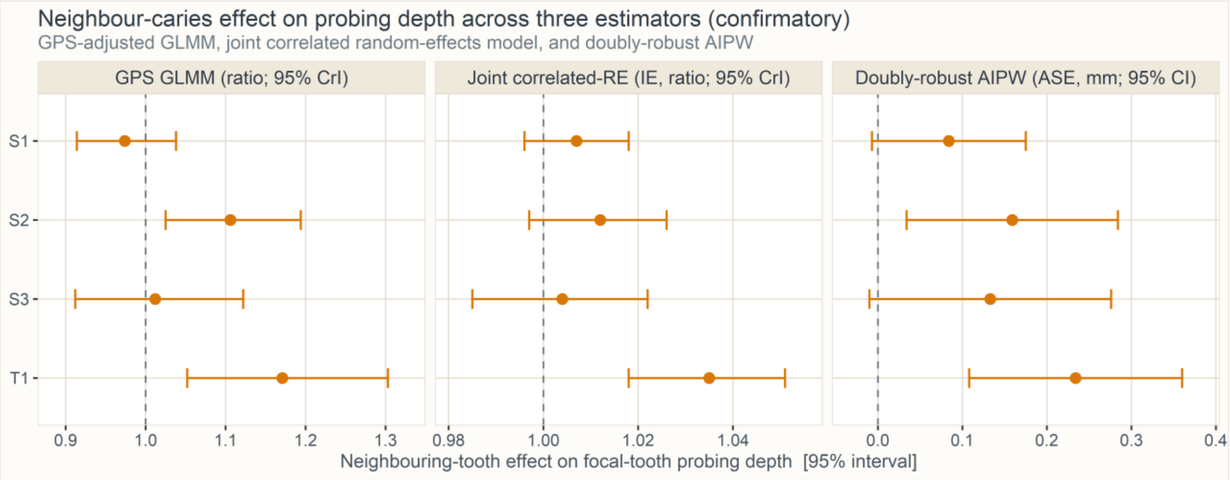
